# Stages of COVID-19 pandemic and paths to herd immunity by vaccination: dynamical model comparing Austria, Luxembourg and Sweden

**DOI:** 10.1101/2020.12.31.20249088

**Authors:** Françoise Kemp, Daniele Proverbio, Atte Aalto, Laurent Mombaerts, Aymeric Fouquier d’Hérouël, Andreas Husch, Christophe Ley, Jorge Gonçalves, Alexander Skupin, Stefano Magni

## Abstract

**Background:** Worldwide more than 72 million people have been infected and 1.6 million died with SARS-CoV-2 by 15th December 2020. Non-pharmaceutical interventions which decrease social interaction have been implemented to reduce the spread of SARS-CoV-2 and to mitigate stress on healthcare systems and prevent deaths. The pandemic has been tackled with disparate strategies by distinct countries resulting in different epidemic dynamics. However, with vaccines now becoming available, the current urgent open question is how the interplay between vaccination strategies and social interaction will shape the pandemic in the next months.

**Methods:** To address this question, we developed an extended Susceptible-Exposed-Infectious-Removed (SEIR) model including social interaction, undetected cases and the progression of patients trough hospitals, intensive care units (ICUs) and death. We calibrated our model to data of Luxembourg, Austria and Sweden, until 15th December 2020. We incorporated the effect of vaccination to investigate under which conditions herd immunity would be achievable in 2021.

**Results:** The model reveals that Sweden has the highest fraction of undetected cases, Luxembourg displays the highest fraction of infected population, and all three countries are far from herd immunity as of December 2020. The model quantifies the level of social interactions, and allows to assess the level which would keep *R*_*eff*_ (*t*) below 1. In December 2020, this level is around 1/3 of what it was before the pandemic for all the three countries. The model allows to estimate the vaccination rate needed for herd immunity and shows that 2700 vaccinations/day are needed in Luxembourg to reach it by mid of April and 45,000 for Austria and Sweden. The model estimates that vaccinating the whole country’s population within 1 year could lead to herd immunity by July in Luxembourg and by August in Austria and Sweden.

**Conclusion:** The model allows to shed light on the dynamics of the epidemics in different waves and countries. Our results emphasize that vaccination will help considerably but not immediately and therefore social measures will remain important for several months before they can be fully alleviated.

## 1 Introduction

In December 2019, a novel strain of coronavirus SARS-CoV-2 (severe acute respiratory syndrome coronavirus 2) was identified in Wuhan (China). In severe cases, it causes an acute respiratory distress syndrome (ARDS), which can lead to respiratory failure, septic shock, multi-organ failure and death [1]. Worldwide 72,318,997 confirmed cases and 1,613,742 dead people have been identified to be infected with SARS-CoV-2 by mid December 2020.

To mitigate the coronavirus disease 2019 (COVID-19) pandemic, mathematical modelling has become a major tool to better understand its spreading [2, 3]. As COVID-19 is currently widely spread across the globe, short- and medium-term modelling forecasts inform the need for containment and mitigation strategies. For these purposes, one of the most widely employed models is the standard Susceptible-Exposed-Infectious-Removed (SEIR) model, for which numerous extensions have been developed during the past months. An extension of the SEIR model has been utilized in [4] to model the COVID-19 epidemic and the implementation of population-wide interventions in Italy. Extensions including compartments for hospitals and dead have been developed e.g. for Italian regions [5] and to predict hospital demand in Colombia [6]. The model presented in [7] developed an extension of the standard SEIR model with hospitals, ICUs and age structure specifically tailored to investigate effects of non-pharmaceutical mitigation in Sweden. SEIR-based models are used to forecast the trajectories of the pandemic and the effect of non-pharmaceutical interventions in the United States [8] as well.

We have previously investigated the importance and impact of various non-pharmaceutical interventions during the early phases of the pandemic [9]. Therein, with an extension of the SEIR model applied to data of several countries, we showed that various synergies of different mitigation strategies could lead to similar results on the suppression of the infection, with potentially different consequences. We also investigated the interplay between epidemiological and economical aspects of the pandemic in Luxembourg in [10]. Here, we further extend the SEIR model including a social interaction parameter, the presence of undetected cases, the disease progression through hospitals, ICU and death and finally vaccination. The aim is to understand the different phases of the pandemic which occurred in several countries that employed different policies, and to investigate how the interplay between vaccination strategies and social interaction might lead us toward herd immunity throughout 2021.

The model is calibrated on available public data collected for Luxembourg, Austria and Sweden. The model was initially developed for Luxembourg, the country with most detected COVID-19 cases per 100,000 inhabitants in Europe as of December 2020. It was further applied to Austria to assess its generality, by proving its ability to capture, with appropriate parameter changes, the evolution of the pandemic in countries exhibiting similar dynamics. We finally applied it to Sweden to assess what additional factors need to be accounted for when describing a country with a very different intervention strategy, which did not impose a full lockdown and is often considered having attempted to reach herd immunity early on. We utilize our model to compare the epidemic dynamics during multiple infection waves within and between each of these countries.

We cross-validated the calibration of our model by means of Bayesian inference and Markov Chain Monte Carlo (MCMC) methods. These methods have been widely applied in the framework of the COVID-19 pandemic to infer epidemiological parameters from SEIR-based or other epidemiological models, e.g. for Spain [11], Germany [12], South Africa [13] and in a comparative study of 11 countries, including Austria and Sweden [14]. Most of these studies employing MCMC were dealing with models not including parameters for hospitalisations, ICU, dead and undetected cases like our present paper, and all of them focus on one country only, except [14] which includes multiple countries but focuses mostly on estimating the basic reproduction number *R*_0_.

Several vaccines are becoming available by the end of 2020 and systematic vaccination campaigns are starting or about to start in several countries. Vaccination aims at protecting others from the infection, as well as the treated individuals [15, 16]. Hence, vaccination is of extreme interest, not only to protect the most vulnerable, but also to contribute to the eradication of the virus in the population through herd immunity. Herd immunity would be achieved when a certain fraction of the total population would become immune to the infectious disease (through recovering from natural infection or through vaccination), so that the infectious agent can no longer generate large outbreaks [17]. Nevertheless, much remains to be learned regarding immunity to SARS-CoV-2 [18]. Moreover, there are several challenges in reaching herd immunity to SARS-CoV-2 infection by mass vaccination, including the supplying of vaccines and the logistic of their deployment to a population. A key question in the current COVID-19 pandemic is thus how and when herd immunity could be achieved and at what cost [17].

To tackle this question, we use our model to investigate the potential impact of various vaccination strategies and their synergy with social measures for the countries considered here. We investigate which conditions on vaccination management would lead to herd immunity by spring and estimate when it could be reached, depending on the various combinations of vaccination strategies and social interaction scenarios. It needs to be stressed that the desirable objective would be to achieve herd immunity primarily by mass vaccination, while avoiding the saturation of healthcare systems and having as few cases and dead as possible. The use of mathematical models in the design of mass immunization programs is a long standing tradition [19], and [20] has investigated the interplay between social interaction and vaccination with a simple Susceptible-Infectious-Recovered-Susceptible (SIRS) model not fit to real countries. Also [21] investigates under which conditions herd immunity could be reached, focusing on vaccination alone and with a simple model that, while including vaccine efficacy and duration of protection, is not fit to data from real countries. Thus, to the best of our knowledge, the model we present here is to date the first one, calibrated on COVID-19 data from multiple countries and including compartments for disease progression and undetected cases, which systematically investigates the interplay of vaccination strategies and plausible social interaction scenarios in the pursuit of herd immunity.

The paper is organized as follows. The mathematical model is introduced in Sec. 2.1, its calibration to available data for the selected countries in Sec. 2.2. Model comparison with data, in order to confirm the validity of our modelling approach, is shown in Sec. 3.1, together with projections for possible future scenarios under different social interaction conditions and an estimate of undetected cases over time. In Sec. 3.2 we present our estimates of social interaction and its effect on the effective reproduction number *R*_*eff*_ (*t*) over time. In Sec. S5.4 we present how data for Sweden can be fit with the same model structure. In Sec. 3.3 we fit subsequent waves of infection with parameter changes in the same model. We further use our model in Sec. 3.4 to investigate the potential effects of vaccination and the interplay between alternative vaccination strategies and social interaction levels. In Sec. 3.5, we estimate when herd immunity might be reached, depending on the vaccination strategy and social interaction level for each country. We discuss these results in Sec. 4 and we draw conclusions in Sec. 5. The detailed mathematical model is described in the supporting material (from Sec. S1), along with the employed data (from Sec. S5) and Bayesian Inference methods (from Sec. S7).

## 2 Model and Methods

### 2.1 Mathematical model

We develop a new mathematical model of the transmission of COVID-19 within a country’s population, extending the standard Susceptible-Exposed-Infectious-Removed (SEIR) model [22] to include 1) undetected cases; 2) varying social interaction; 3) the progression of severe cases through hospitalization, intensive care and eventually death or recovery and 4) vaccination.

The model, whose structure is depicted in Fig. 1, is implemented through a set of ordinary differential equations Eq. (S2). The total population *N* of the considered country is streamed at time *t* into 16 compartmental variables, summarized in Tab. S1. Each variable represents the number of individuals at a particular stage, normalized by the total country population. The parameters of the model represent either probabilities *p*_*i*_ of going to one compartment or another (with *i* being an index over parameters, specified in Tab. S2), or rates *τ*_*i*_ describing how fast individuals flow through compartments. The parameter *ρ* (*t*) is a tuning for the average contact rate *β*. These parameters are detailed in Tab. S2.

**Fig. 1.**
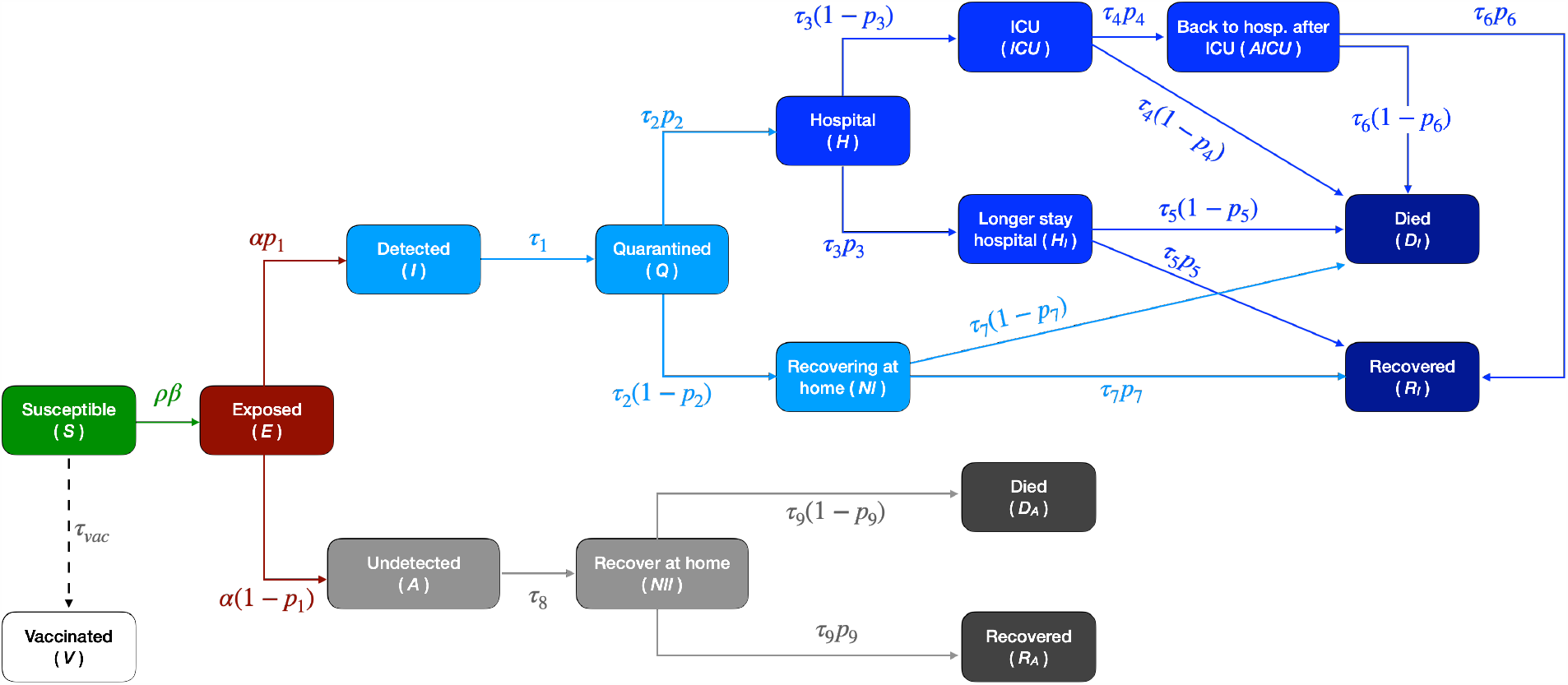
Scheme of the mathematical model. Each compartment is associated to a variable (*cf*. Tab. S1 and Eq. (S2)). Each variable represents the fraction of individuals being in that state at a given time; arrows represent the flow of individuals between states, with associated probabilities and rates (Tab. S2). Their values (for each country and wave) are summarized in Tab. S6.

An important fraction of infected people is usually not detected [23], so we introduced a separation between detected and undetected cases. One branch (detected cases) can lead to quarantine and possibly to hospital and/or ICU. The one of undetected cases (either because they are asymptomatic or simply due to lack of testing) continues spreading the infection until recovery or death. The probability of an infected individual to be detected is indicated as *p*_1_ and assumes different values for each country and wave of infections, *cf*. Tab. S2 and Tab. S6.

The population flow from one compartment to another is governed by rates multiplied by probabilities. While the use of rates is inherited from the SEIR model, probabilities are not employed therein because in the SEIR individuals can follow only one path. Probabilities determine which fraction of the population goes to one branch, which to the other. Rates, representing the inverse of the average length of stay in a compartment, capture how fast individuals flow from one pool to the next (on average).

The parameter *ρ* (*t*) represents the fraction of social interactions taking place at any given time, with respect to the one pre-pandemic. Thus, a value of *ρ* (*t*) = 1 means the same level of social interaction than in the beginning of 2020. This parameter incorporates all non-pharmaceutical interventions and changes in population behaviour that affect the dynamics in a similar manner. It changes whenever new measures or major updates in social interactions take place (lockdown, schools opening/closure, summer vacation, etc.) and is further discussed in Sec. 3.2.

We include additional compartments to model the different phases of the disease for infected individuals. In particular, we consider the progression to more severe states, requiring hospitalisation and intensive care (ICU), and eventually death in a fraction of cases.

### 2.2 Model calibration and fit to public data

The model is fitted as described in Sec. S6 to available public time-series data (from February to 15th December 2020), separately for each wave of infection and for each of the countries mentioned in the introduction: Luxembourg (data and dates discussed in Sec. S5.1), Austria (Sec. S5.2) and Sweden (Sec. S5.3). These data are displayed in Fig. 2 for total cases (**A, B, C**) and corresponding daily new cases (**D, E, F**), hospital occupation (**G, H, I**), ICU occupation (**J, K, L**) and deaths (**M, N, O**). Subsequent waves of infections have occurred; we refer to them as 1st and 2nd wave for Austria and Sweden, respectively for the ones starting in February and September. In Fig. 2 **D**, we notice that Luxembourg had an additional wave during July, which appears distinct from the one in March and the one in September (though this separation is somewhat arbitrary). So, we refer to it as 2nd wave, and we will refer to the rise in September as 3rd wave.

**Fig. 2.**
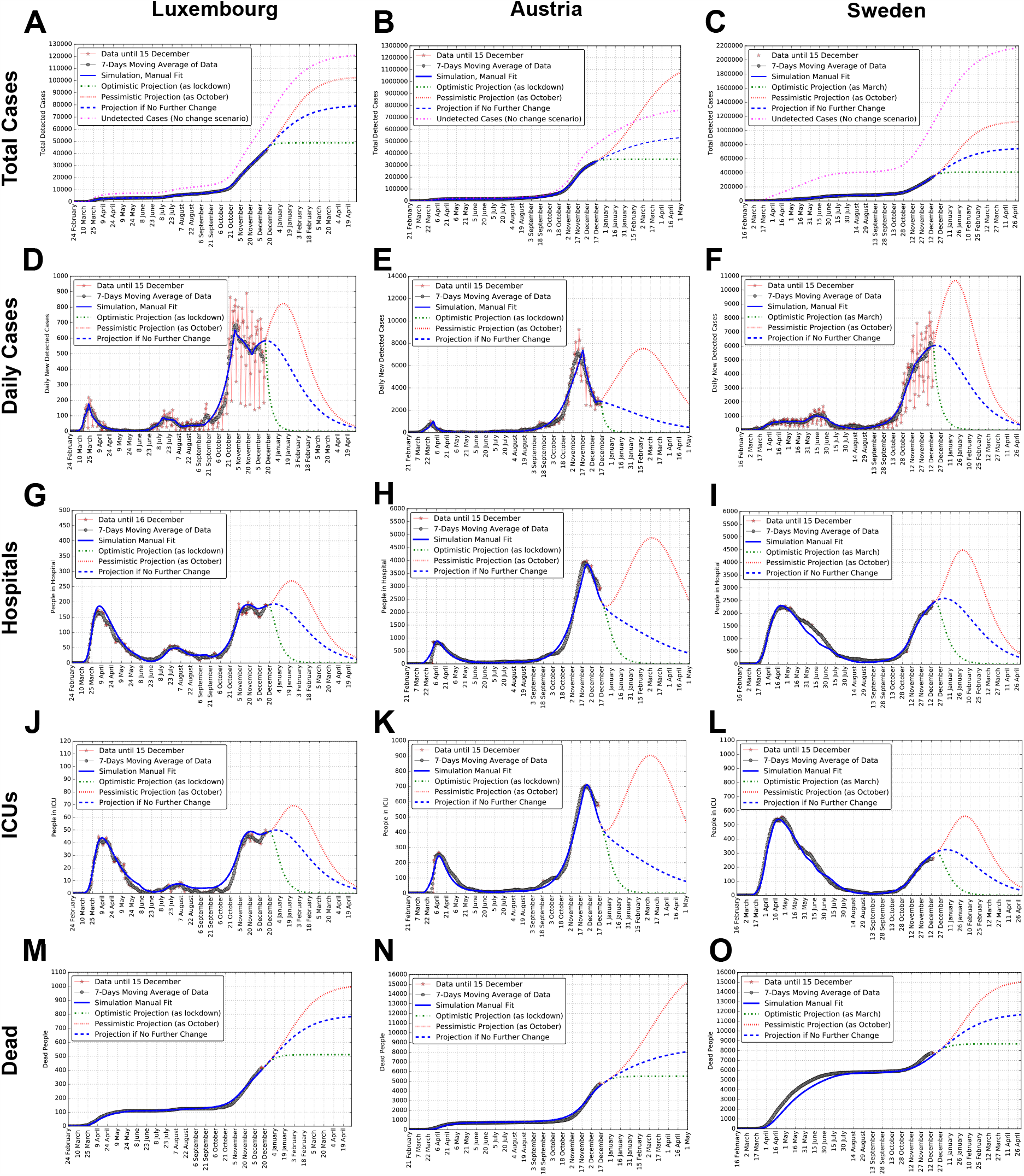
Data, model simulations and projections of total cases, daily cases, hospital and ICU occupation and dead, for each country. Luxembourg (**A, D, G, J, M**), Austria (**B, E, H, K, N**) and Sweden (**C, F, I, L, O**). Total cases (**A, B, C**) and daily new cases (**D, E, F**) from Eq. (S8). Hospital occupation (**G, H, I**) from Eq. (S9). ICU occupation (**J, K, L**) from Eq. (S10). Dead (**M, N, O**) representing the sum of Eq. (S11) and Eq. (S12). In addition to model simulation, raw data are shown, together with their weekly moving average. Panels **A, B, C** also report the cumulative number over time of undetected cases in the model, obtained as the sum of the variables *A* + *NII* + *D*_*A*_ + *R*_*A*_, in violet. Projections for several months after the last data point illustrate potential simulated scenarios, corresponding to no change in social interaction, an optimistic scenario where the social interaction parameter is as low as during lockdown/March, and a pessimistic scenario where the social interaction parameter is as high as in October.

For each country and each epidemic wave, we initially calibrated manually a set of parameters that allow the model simulations to fit the data (*cf*. Sec. S6). Rates and probabilities used are listed in Tab. S6 and the values of the social interaction parameter, and its changes over time, are displayed in Fig. 3 and listed in Tab. S3 for Luxembourg, Tab. S4 for Austria and Tab. S5 for Sweden. Model simulations based on these parameters are shown in Fig. 2, where we can appreciate that our country-specific models fit remarkably well the weekly moving average of data. This supports our choice of parameters, as for each country and wave our model is able to fit simultaneously several time-series data.

**Fig. 3.**
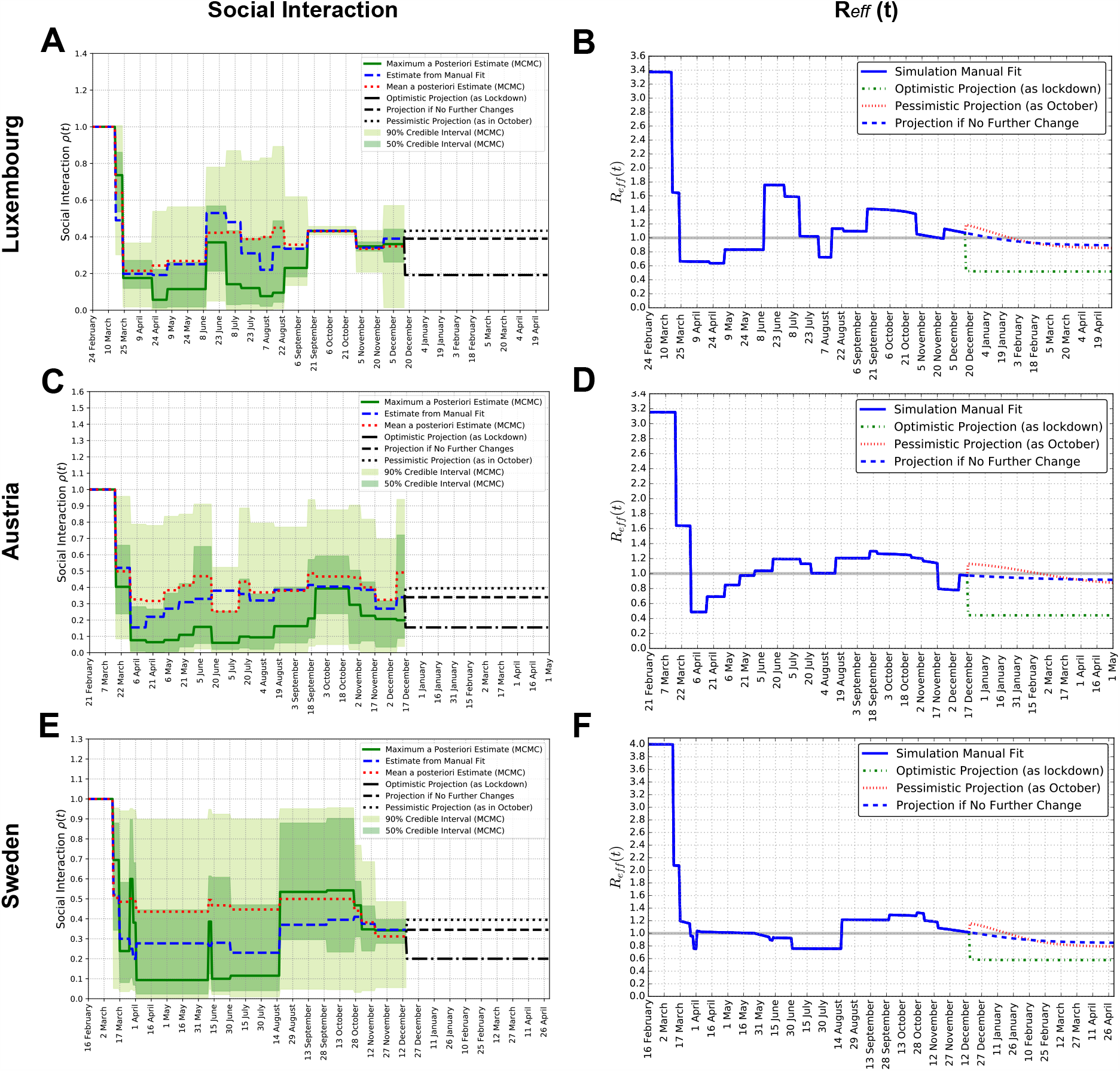
Social interaction parameter *ρ* (t) and effective reproduction number *R*_*eff*_ (t), for each country. Panels **A**,**C**,**E**: Social interaction parameter *ρ* (*t*), manual and MCMC fits for Luxembourg, Austria and Sweden respectively. Dates of change and parameter values are reported in Tab. S3 for Luxembourg, Tab. S4 for Austria and Tab. S5 for Sweden. Panels **B, D, F**: Effective reproduction number *R*_*eff*_ (*t*), computed from Eq. (S6) analytically derived in Sec. S4, corresponding to the values of the social interaction parameter for Luxembourg, Austria and Sweden, from their manually calibrated parameter sets.

We further cross-validated the calibration of our models by means of Bayesian inference and Markov Chain Monte Carlo (MCMC) methods in Sec. S7. A set of MCMC simulations was performed to obtain, within a Bayesian framework, an estimate of the parameters of our model and to quantify their uncertainties. The choice of prior probability distributions for the parameters is specified in Sec. S7.2 and the convergence of the MCMC chains is verified in Sec. S7.5. As we can see from Sec. S8.1, Fig. S2, the values of the manual fit are usually included within the Bayesian credible intervals, which are rather wide, thus the manually calibrated parameter sets are consistent with the Bayesian estimates.

We observe in Sec. S8.2, Fig. S3, that there is high degeneracy between parameters, with combinations (e.g. ratios) of parameters being better constrained than the individual parameters. The MCMC is extremely useful to estimate the uncertainty that affects our estimate of parameters and their potential ranges of values. Nevertheless, it does not provide a unique parameter set due to poor identifiability, common to SEIR models [24]: the two parameter sets obtained respectively from the maxima of the posteriors (maximum a posteriori estimate, MAP) and from the means of the posteriors are rather different from each other due to asymmetric posteriors. Moreover, certain parameters are poorly constrained by the data, resulting in rather flat posterior probability distributions. For the simulations, we thus employed the manually calibrated parameter set, which is unique, consistent with the results of the MCMC as explained above, and incorporates domain knowledge from literature.

### 2.3 Time-dependent probabilities of detection and hospitalisation for Sweden

In order to investigate the model properties, we applied it to Luxembourg and Austria, which implemented similar strategies resulting in similar epidemic dynamics, and furthermore for comparison we apply it to Sweden, which choose very different strategy and thus experienced a different epidemic dynamics. We then compare the simulations and the model-building process among the three countries. To fit Swedish data with the same model structure as above, it was not sufficient to consider a different set of parameter values. Instead, it was necessary to introduce time-dependent probabilities of being detected *p*_1Swe_ (*t*) and of being hospitalized *p*_2Swe_ (*t*), within an epidemic wave. These are derived in Sec. S5.4 and are depicted in Fig. S1.

We estimated *p*_1_, the probability of an infected individual being detected, from prevalence data [25, 26]. While the particular functional form chosen for *p*_1*Swe*_ (*t*) is somewhat arbitrary and multiple forms could be employed (*cf*. Sec. S5.4), for Sweden a time-dependent *p*_1*Swe*_ (*t*) increasing with time seems indeed necessary in order for the model to fit the available data, which was not the case for Luxembourg and Austria, where *p*_1_ and *p*_2_ values constant within each wave were sufficient.

We further need to incorporate in the model the observation that during the early phases of the pandemic in Sweden the majority of the detected cases were people needing hospitalization. This can be clearly seen from Fig. S1, panel **C**, which shows data for daily new detected cases, people entering hospital and people entering ICU (not to be confused with the number of people currently occupying hospitals and ICUs) publicly available on [27] and ^1^. It becomes obvious that in March and April the daily number of people entering hospital was more than half of the daily number of detected cases, leading us to employ a time-dependent *p*_2*Swe*_ (*t*). This clearly changed in May and June, likely due to the change in testing strategy ordered by the government on May the 3rd [28].

## 3 Results

### 3.1 The model accounts for undetected cases and provides future projections of potential scenarios

We use our model to simulate potential future developments of the epidemic in each country. While very instructive, it should be stressed that projections might be accurate for few weeks, but much less on longer time periods, because small changes in e.g social interactions and corresponding parameters can lead to large changes in the simulated outcome. Consequently, their main goal is to inquire potential scenarios that might unfold from the current situation [11]. In Fig. 2, we simulate the potential evolution of the epidemic in the coming months until spring. After the last data-point, we simulate three scenarios, to illustrate alternative potential outcomes, namely one corresponding to no change in social interaction, an optimistic scenario where the social interaction parameter becomes as low as during the lockdown in March, and a pessimistic scenario where it becomes as high as in October. These three scenarios are meant to represent an average outcome and two extreme but plausible cases.

Such projections are sensitive to the fraction of the population that has already been infected, but has not been detected. Our model includes compartments to describe that fraction, which is mostly controlled by the probability *p*_1_ of being detected. This was estimated for each country and wave based on the available prevalence data, as discussed in Sec. S6 and reported in Tab. S6. Panels **A, B** and **C** also report the cumulative number of undetected cases in the model, obtained as the sum of the variables *A* + *NII* + *D*_*A*_ + *R*_*A*_, for each country. We can see that, over time, this number (violet dashed curve) is approximately between one and two times the number of detected cases for Luxembourg and Austria, while it is up to three to four times for Sweden. Nevertheless, for Austria and Sweden the sum of the number of detected cases and the estimated number of undetected cases until December remains more than an order of magnitude smaller than the population of the country, while for Luxembourg it is higher. In fact, our model estimates that until the 15th December, the percentage of the population having been infected by SARS-COV-2 in Luxembourg is about 18.3% (7.2% detected and 11.1% undetected); in Austria 9% (3.7% detected and 5.3% undetected) and in Sweden 14.5% (3.5% detected and 11% undetected). For all three countries, this makes herd immunity still far from being reached.

### 3.2 Social interaction drive epidemic dynamics and *R*_*eff*_ (*t*), across countries

In this section, we present our estimates of the social interaction parameter *ρ* (*t*), and how it affects the effective reproduction number *R*_*eff*_ (*t*) over time, for Luxembourg, Austria and Sweden.

The parameter *ρ* (*t*) tunes social interactions, and can evolve when new measures or major behavioural changes occur. Hence, it is implemented as a piece-wise constant function. Its value changes at specific dates *t*_*n*_ when modifications in policies took place. We assume a mean constant value *ρ*_*n*_ over the subsequent period of time. For the three countries considered, measures, dates and values of *ρ*_*n*_ are summarized in Tab. S3 for Luxembourg, Tab. S4 for Austria and Tab. S5 for Sweden. The evolution of *ρ*_*n*_ is depicted in Fig. 3 respectively for Luxembourg (panel **A**), Austria (panel **C**) and Sweden (panel **E**), for both the manually calibrated parameter set and the Bayesian inference results.

These panels show values of *ρ*_*n*_ from manual calibration (Sec. S6) and from Bayesian inference (Sec. S8.1 and Fig. S2), which are proportional to the number of daily cases (*cf*. Fig. 2). Our manually calibrated social interaction values are consistent with our Bayesian estimates, as they are usually very close to the mean or the maximum of the Bayesian estimate, or fall within the 50% credible interval (dark green bands), or the 90% (light green bands). This supports the validity of the social interaction values from our manually calibrated parameter set, which was obtained by requiring that model simulations fit reasonably well the moving average of the data of Fig. 2, panels **A, B, C**. Changes in the social interaction parameter lead to a very good fit of the model to the data of detected cases. This identifies the social interaction as an essential parameter for the model. In turn, this underlines the importance of social interaction management for epidemic control.

Next, we investigated the dependency of the effective reproduction number *R*_*eff*_ (*t*) on the social interaction parameter and on the fraction of population still being susceptible. The time evolution of *R*_*eff*_ (*t*) obtained by Eq. (S6) (*cf*. Sec. S4) for each country is displayed in Fig. 3, panels **B, D** and **F**. Step-wise changes in *R*_*eff*_ (*t*) arise from changes of *ρ* (*t*), while gradual changes in *R*_*eff*_ (*t*) are instead associated to the depletion of the pool of susceptible individuals.

The value of *R*_*eff*_ (*t*) at the beginning of the epidemic provides the basic reproduction number *R*_0_ ≐ *R*_*eff*_ (*t* = 0). We obtain estimates of 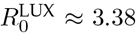 for Luxembourg, 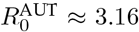 for Austria and 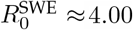 for Sweden. Such values can be compared with those obtained from independent methods. 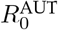 and 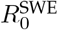 are consistent with those obtained from previous Bayesian estimates [14], within credible intervals. In addition, the complete time evolution of 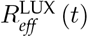 is consistent with what is reported on the official Luxembourg Government website ^2^, which is assessed with independent methods. Panels **B, D** and **F** of Fig. 3 depict *R*_*eff*_ (*t*) evolution over time, for each country.

Finally, from our estimate of *R*_*eff*_ (*t*) depicted in panels **B, D** and **F** we can evince the value of *R*_*eff*_ at the date of the last data-point, namely 15th of December, which we find to be 1.07 for Luxembourg, 0.97 for Austria and 1.01 for Sweden. Interestingly, all three countries seems to be, as of mid December, very close to *R*_*eff*_ ≈ 1. We further estimate the percentage of social interaction that is currently needed to have *R*_*eff*_ (*t*) ≈ 1; this is similar across countries: 36% (w.r.t. before the pandemic and corresponding measures) in Luxembourg and 34% in both Austria and Sweden.

### 3.3 Parameter fitting reveals probability of hospitalization decreases between waves

The social interaction parameter *ρ* (*t*) changes a limited number of times to represent changes in policies or population behaviour. To fit the first waves of Luxembourg and Austria, all other parameters are constant (though somewhat different between the two countries). This results from the fact that the time evolution of daily new cases, hospital and ICU occupation are alike in these two countries, although scaled and delayed. However, the same parameters from the first wave overestimate the hospital pro-gression during the second wave.

Fig. 4 reports the parameter changes between one wave and the subsequent, in terms of the log-2 of the fold change, i.e. *log*_2_ (*FC*), with 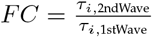. Thus, *FC* = 0 means no change, *FC* = 1 means that the parameter in the second wave is twice as big as in the first wave, *FC* = − 1 means that the parameter in the second wave is half of its value in the first wave, and so on.

**Fig. 4.**
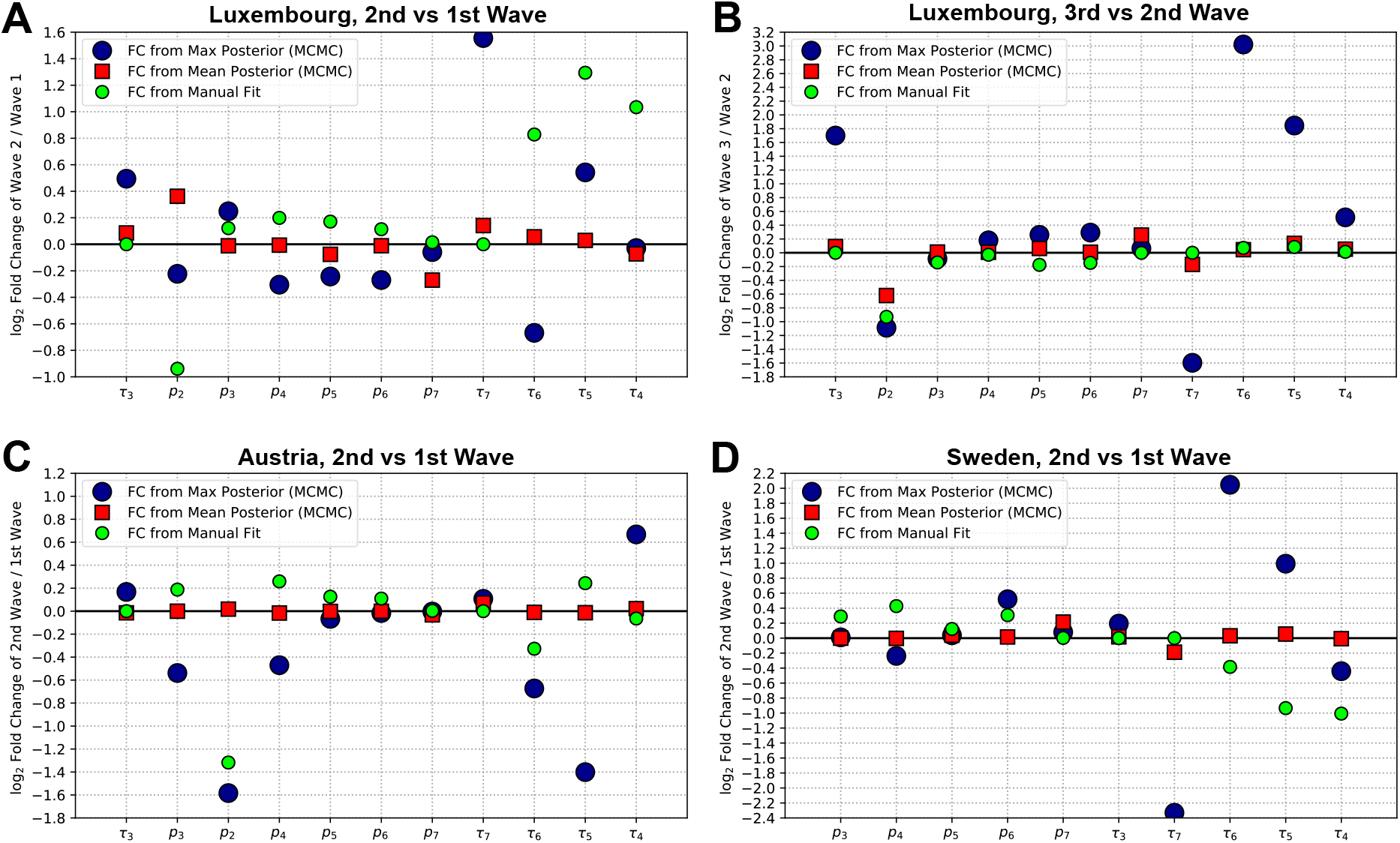
Fold-changes (FC) of the point-estimates of each parameter, between a wave and the subsequent, for each country. **A**: 2nd wave versus 1st wave, Luxembourg. **B**: 3rd wave versus 2nd wave, Luxembourg. **C**: 2nd wave versus 1st wave, Austria. **D**: 2nd wave versus 1st wave, Sweden. The values for the parameters employed are displayed in Fig. S2 and summarized in Tab. S6 for the manual fit. The vertical axes indicate log_2_ (*FC*) for each parameter. The green small dots depict the fold changes for the manually calibrated parameter sets. Errors on the means were computed by the standard error of the mean, but are too small to be noticeable in the figure.

As we can see, in the manual fit parameter sets (green circles) we considerably decreased *p*_2_, the probability of being hospitalized (when detected positive) between each wave and the subsequent, both in Luxembourg and in Austria (together with other parameter changes). We also compute the fold-changes arising from the two sets of parameters obtained by Bayesian inference, namely the one from the maxima of the posterior probability distributions, and the one from their means, and we display them in Fig. 4. We see that the parameter *p*_2_ decreases considerably also in the maximum a posteriori estimate (MAP, blue dots) from the MCMC both between the 2nd and 3rd wave of Luxembourg (panel **B**), and between the 1st and 2nd wave of Austria (panel **C**). Only from the 1st to 2nd wave of Luxembourg the MCMC shows a slight decrease for the maximum of the *p*_2_ estimate, but conversely a slight increase in the mean. A large decrease in *p*_2_ is also what we assume in Sweden, where as introduced in Sec. 2.3, detailed in Sec. S5.4 and displayed in Fig. S1 panel **D**, we assume (based on data of new daily cases and hospital admissions) a decrease in *p*_2_ (*t*) from 0.9 in March to 0.1 from June onward. These findings altogether underline the fact that, overall, the probability of being hospitalized when detected (*p*_2_) decreased for subsequent waves of a country, and this for all three countries investigated.

More in general, the means supposedly provide more robust information than the maxima. It should be reminded that the maxima are representative of the posterior distribution only when the posterior has a clear peak and is not almost flat, otherwise the maxima are not particularly representative. This leads to some of the FCs obtained from the maxima (large blue dots) having rather extreme values, in particular those associated with rates *τ*_4_, *τ*_5_, *τ*_6_ and *τ*_7_, the rates involved in the patient progression through hospital and ICU, as depicted in Fig. 1. Instead, as we can see in the figure, the fold changes computed employing the means tend to be very often close to 0, meaning no significant change in the posterior mean for that parameter between subsequent waves.

The MCMC estimate does not fully reflect the changes in all parameters that we performed between one wave and the next in the manually calibrate parameter set (*cf*. Tab. S6). We interpret these discrepancies in the fold-change for few parameters between manual fit and MCMC parameters sets (*cf*. Fig. 4) as indicating that, due to the large uncertainty in the identification of the parameter values which our Bayesian inference approach underlined, multiple parameter sets could provide an equally good fit of the model to the data.

### 3.4 Simulations show that social measures still have significant impact on the infection curve in early 2021, along with vaccination

The model is extended as summarized in Sec. S3 to include fully protective vaccination and to investigate the interplay between its dynamics and social interaction. Three potential vaccination strategies starting from 1st January 2021 are simulated: vaccinating all the population of the country within the first 6 months of 2021, within one year, or within 1.5 years. These represent three strategies among the potential timelines that various countries might attempt to implement. For the countries considered, 6 months could represent the scenario of a very fast vaccination, 1 year an average assumption and 1.5 years could correspond to envision a slow process of vaccination of the population. We assume that the vaccination capability scales with country size (*cf*. Sec. S3). To compare future projections, we also consider the baseline case where no vaccination is performed. The results are displayed in Fig. 5. Panels **A, B, C** report the time evolution of the number of vaccinated people for the various scenarios and strategies, panels **D, E, F** display *R*_*eff*_ (*t*) and panels **K, L** and **M** the number of total detected cases. Dashed curves symbolize the four different vaccination strategies. The objective being to investigate the interplay between social contacts and vaccination strategies, we report the curves corresponding to combinations of the two. We consider two alternative values of the social interaction parameter *ρ*: a value corresponding to the pessimistic scenario of Fig. 2 (which corresponds to going back to the social interaction level of October) in red, and a value corresponding to the average scenario of Fig. 2 (which corresponds to no changes in the social interaction w.r.t. the value of November) in blue. The optimistic scenario for social interaction is not included because it corresponds to a full lockdown, unlikely to be applied for several months consecutively, and for visual clarity.

**Fig. 5.**
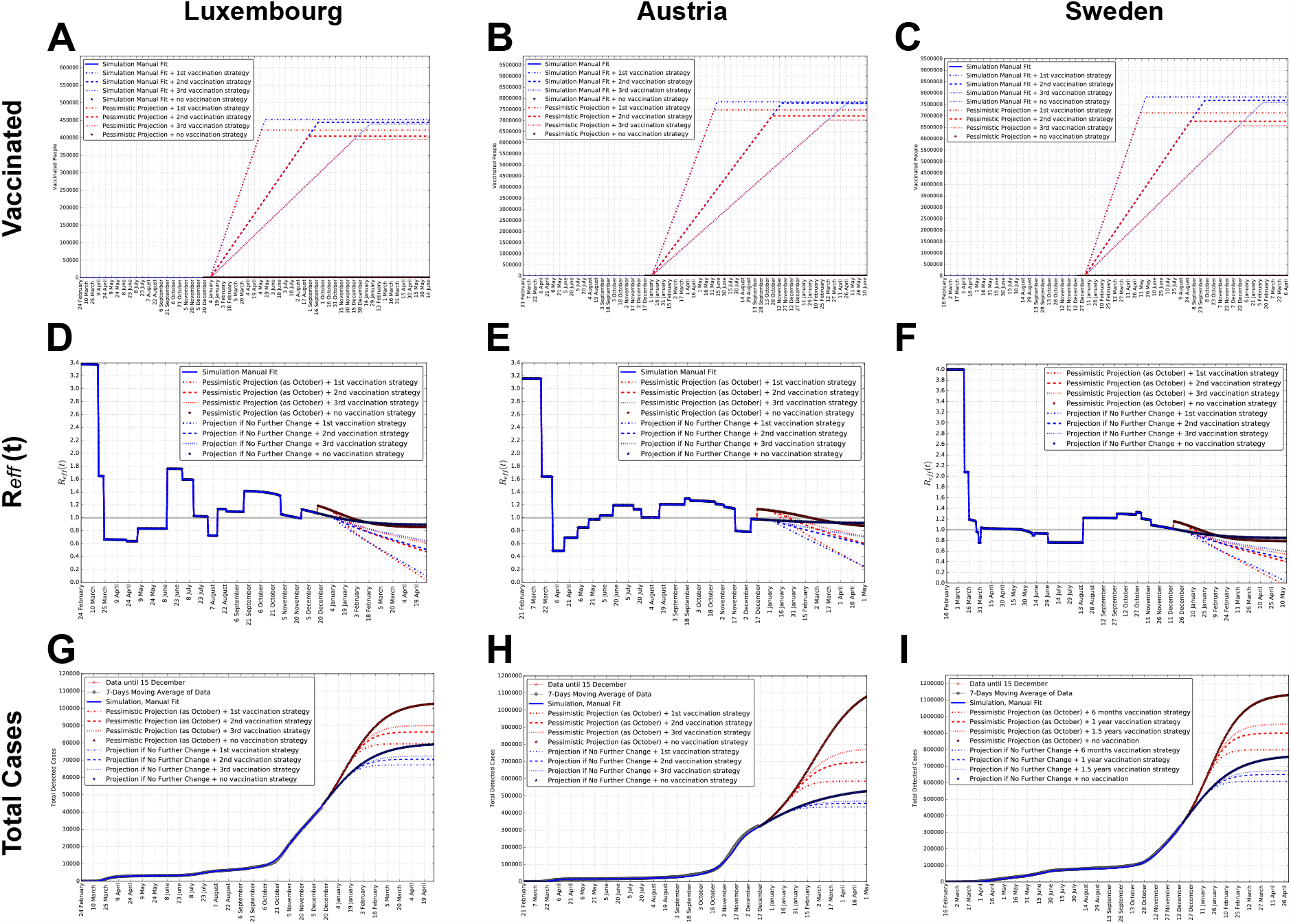
Simulations of different vaccination scenarios: projections of number of vaccinated people, *R*_*eff*_ (*t*) and total detected cases for each country. Luxembourg (**A, D, G**), Austria (**B, E, H**) and Sweden (**C, F, I**) are reported. Projections for several months after the last data point illustrate potential simulated scenarios, for three alternative vaccination strategies and for no vaccination, both in the case of the “pessimistic” scenario (red, corresponding to social interaction as high as in October) and the “no change” scenario (blue, corresponding to no change in the social interaction) from Fig. 2 and Fig. 3. For each social interaction scenario, we show the curves for no vaccination and for 3 alternative vaccination strategies, which correspond to vaccinating all the population of a country in, respectively, 6 months, 1 year or 1.5 years, starting from January 1st, 2021. Panels **A, B** and **C** show the number of vaccinated people as a function of time. Panels **D, E** and **F** show *R*_*eff*_ (*t*). Panels **G, H** and **I** show the number of total detected cases.

Panels **A, B** and **C** in Fig. 5 demonstrate that, for each vaccination strategy, a smaller number of people will be left to be vaccinated if the social interaction parameter is higher, since more people already got infected naturally. We also appreciate that slower vaccination strategies will result in less people to be vaccinated as more people will by then have acquired immunity naturally. These observations are consistent across the three countries considered.

Panels **D, E** and **F** report the simultaneous effects of social interaction and of the shrinking of the pool of susceptible, due to both infection and vaccination, on *R*_*eff*_ (*t*). In each country, for any vaccination strategy as well as for no vaccination, the pessimistic scenario (red) starts with a higher *R*_*eff*_ (*t*) than the “no change” scenario (blue). However, the situation is soon inverted due to the larger shrinking of the susceptible pool associated with a higher social interaction parameter. This suggests that the interplay between the two measures (social interaction and vaccination) is non-trivial, and will require deeper investigation, which we will perform in the next section.

Panels **G, H** and **I** illustrate the changes in the dynamics of total detected cases for the same scenarios and strategies. We observe that the faster the vaccination strategy, the smaller the number of total detected cases reached asymptotically. We also remark that any vaccination strategy (vaccinating all population in 6 months, 1 year or 1.5 years) lead to a considerable reduction of the number of detected cases w.r.t. the case of no vaccination, larger than the difference in cases between the three strategies.

On the other hand, for all three countries the number of total cases is impacted more, or with similar magnitude, by the different social interaction scenarios (red and blue curves) than by vaccination (different dashing for the same color of the curve).

Overall, panels **G, H** and **I** indicate that, on the short term until spring 2021, vaccination will be important to protect the lives of the most vulnerable, but it is going to impact the curve of infections less or as much as the social interaction, depending on the evolution of the epidemic. Thus, social interaction still remains crucial for the time being. Both vaccination strategies and social interaction play a significant role of comparable magnitude to suppress the infection wave. Notably, social interaction could still play a dominant role depending on the context. This triggers the following questions: which combinations of social interaction and vaccination strategy would be needed to achieve herd immunity already e.g. in spring? More broadly, when might herd immunity be reached in the different scenarios? To tackle these questions we further investigate in the next section how will the dynamics of social interaction and vaccination interplay, in a more systematic and complete manner.

### 3.5 Simulations suggest that vaccinating whole population in a year could provide herd immunity not before mid summer

Herd immunity means that, when a certain fraction of the total population become immune to an infectious disease (through recovering from natural infection or through vaccination), the infectious agent can no longer generate large outbreaks [17]. The population fraction that needs to be immune, in order to reach herd immunity in the absence of control measures, is *p*_*c*_ = 1 − 1*/R*_0_ [17]. To answer the question “which combinations of social interaction and vaccination speed would be needed to achieve herd immunity already in April?”, we first compute this quantity for the three countries we investigate. Using the values of *R*_0_ previously estimated in Sec. 3.2, we obtain: *p*_*c,Lux*_ = 1− 1*/*3.38 ≈ 0.70 = 70% for Luxembourg, *p*_*c,Aus*_ = 1 −1*/*3.16 ≈ 0.68 = 68% for Austria and *p*_*c,Swe*_ = 1− 1*/*4.00 ≈ 0.75 = 75% for Sweden. These values are plausible, because similar to the fraction of people which needed to get infected before the epidemic stopped supposedly by naturally achieved herd immunity in Manaus, capital of the Brazilian Amazon. There, 76% of the population was estimated having antibodies in October [29], as the result of a largely unmitigated COVID-19 epidemic, with catastrophic losses of lives. Our estimates means that the pool of susceptible individuals needs to shrink to, respectively for the three countries considered here, 30%, 32% and 25% of the total population. This can be achieved either by vaccination or by surviving natural infection. A reasonable goal should be to achieve this objective as much as possible by vaccination, minimizing the natural infection [30] which can lead to disease and, in a fraction of the cases, to death. Among other reasons [17], this signify that vaccination presents the safest way to reach herd immunity.

In Fig. 6, we display the percentage of the country’s population who will still be susceptible to the virus in April, for all possible values of the social interaction parameter from 16th December on, and of the vaccination speed (assuming vaccination would start on January the 1st), i.e. how many vaccinations per day will be performed. By requiring the percentage of people still susceptible by that date to be equal to or smaller than the fraction *p*_*c*_ needed for herd immunity, we can identify the combinations of parameters for which herd immunity will have already been reached by then. This is shown in the panels by the dashed black areas. Herd immunity can be either reached purely by natural infection (by moving along the vertical axis), or purely by vaccination (by moving along the horizontal axes), or by a combination of the two. This is the most plausible scenario, because for each of the countries presented here the social interaction parameter during the various phases of the pandemic was never 0 and assumed values approximately between 0.05 and 0.5 (*cf*. Fig. 3), corresponding to a reduction of social activity of 5% to 50% w.r.t. before the pandemic. The three vertical lines in the panels of Fig. 6 show the number of vaccinations per day required to immunize the country’s whole population respectively within 6 months, 1 year or 1.5 years, which are computed in Sec. S3. From Fig. 6, panels **A, B** and **C** we can see that vaccinating a country’s population within 1 year at a constant rate could lead to herd immunity by July in Luxembourg and by August in Austria and Sweden. In order to reach herd immunity by mid April primarily by vaccination, we see that Luxembourg needs approximately 2700 vaccinations/day, Austria and Sweden 45000.

**Fig. 6.**
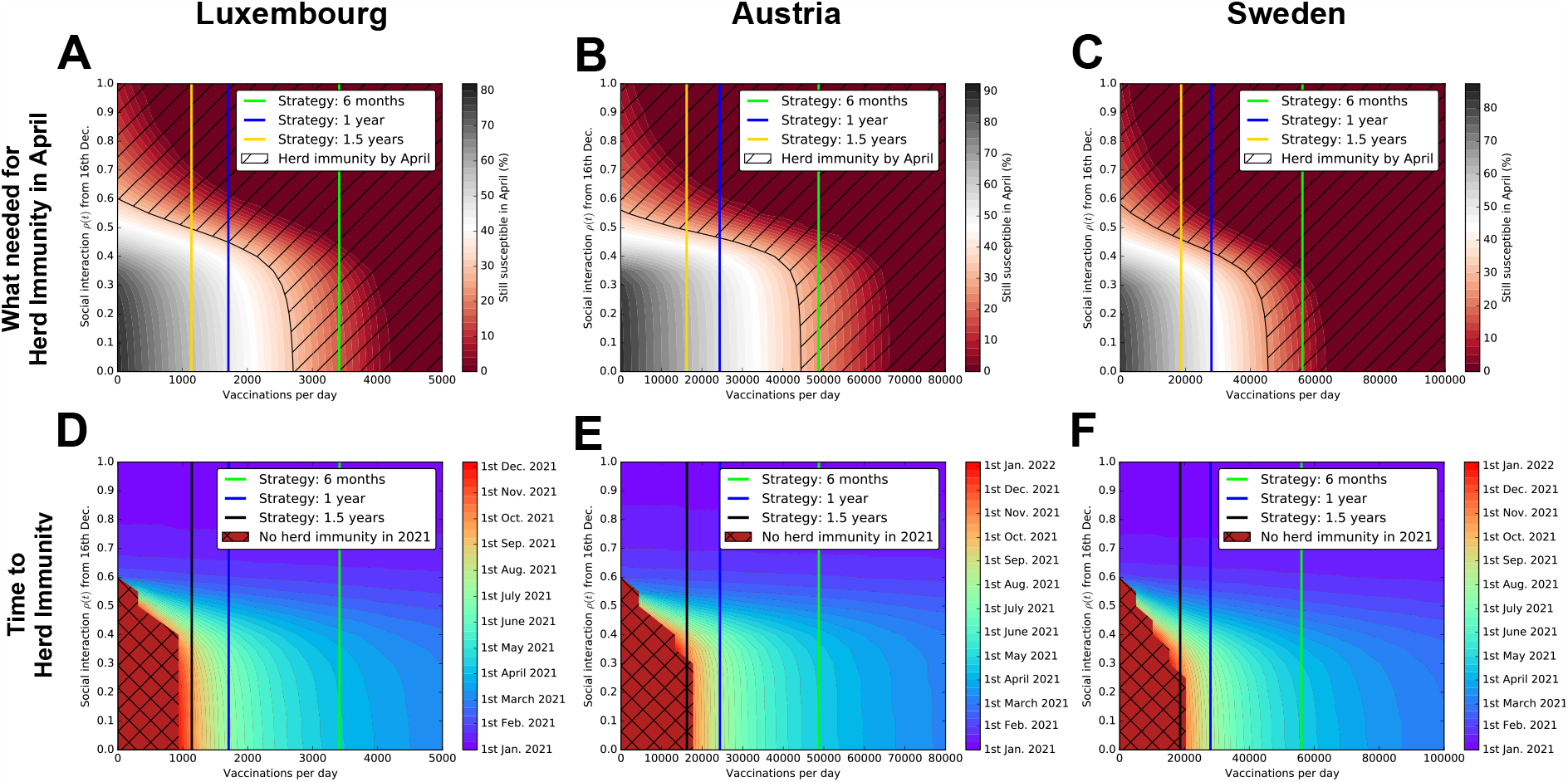
Systematic investigation of the interplay between vaccination strategies and social interaction scenarios, to estimate the time by which herd immunity might be reached. Luxembourg (**A, D**), Austria (**B, E**) and Sweden (**C, F**) are reported. Panels **A, B** and **C** report the number of people who will still be susceptible by April 2021, for all relevant combinations of the social interaction parameter *ρ* and of the number of vaccinations performed per day. The area covered with a black dashing identifies the combinations of social interaction and vaccination speed that would lead to herd immunity by April. We indicate by vertical lines the three alternative vaccination strategies considered in Sec. 3.4, which correspond to vaccinating the whole population of a country in, respectively, 6 months, 1 year or 1.5 years, starting from January 1st, 2021. Panels **D, E** and **F** report the time at which herd immunity might be reached, depending on the combinations of the social interaction parameter *ρ* and of the number of vaccinations performed per day. These dates have to be considered as indicative, since the parameters employed are subject to uncertainty and changes. We indicate with a black grid the combinations of social interaction and vaccination speed that would not allow to achieve herd immunity until 2022. Vertical lines indicate the three vaccination strategies considered.

A broader question we tackle next is: when might herd immunity be reached in the different scenarios? To estimate this, we simulate the time-evolution of our model for all combinations of social interaction parameter and number of vaccinations within the values of interest. We then assess, for each combination of values, at which time the fraction of susceptible population reaches the above mentioned values necessary for herd immunity. We display in Fig. 6, panels **D, E** and **F** the dates at which these thresholds for herd immunity are reached in the corresponding simulations. Such dates are indicative and depend on the assumptions of the present model. Thus, they provide an estimate rather than a prediction. The black grid in Fig. 6 highlights the ranges of parameter values for which herd immunity is not reached within 2021. The three vaccination strategies considered are reported on the plots with vertical lines. We see that without vaccination herd immunity cannot be reached within 2021 with the typical levels of social interaction which we observed so far in any of the three countries considered (usually below, or considerably below, 0.5).

For Luxembourg, all three vaccination strategies might well reach herd immunity within 2021, for Austria and Sweden the 1.5 years strategy would be borderline insufficient, while the other two strategies would do it for each country. If herd immunity had to be reached purely by vaccination, the 6 months strategy would take approximately until April for Luxembourg, and May for Austria and Sweden, and the 1 year strategy until July for Luxembourg and August for Austria and Sweden. Higher (undesired) values of social interaction might anticipate it, but with their undesired consequences. For reference, the assumed vaccination would start on January the 1st, 2021.

Several factors might influence these estimates one way or another. For instance, we assumed reinfections are negligible. Cases of re-infection have been documented [31], if widespread this would create an additional challenge for achieving herd immunity. Nevertheless, re-infection by SARS-Cov-2 has only been conclusively documented in a very limited number of cases so far and it is unclear whether this is a rare or common phenomenon [17]. We extensively discuss it in Sec. 4.7. On the other hand, a recent study [32] based on a model including age distribution has found that the percentage of population necessary to achieve herd immunity might be considerably lower than the typical values proposed, thus making herd immunity potentially less difficult to reach.

## 4 Discussion

### 4.1 Model projections

We have developed a mathematical model extending the standard SEIR model to incorporate 1) the impact of social interaction, 2) the presence of a non-negligible quantity of undetected cases, 3) the progression of the disease for infected individuals and 4) the effects of vaccination. Our model allows us to create projections by simulating potential scenarios and their evolution in the time to come.

For our projections in Fig. 2, the potential evolution of the epidemic is simulated in three alternative scenarios, namely one corresponding to no change in social interaction, an optimistic scenario where the social interaction parameter is as low as during lockdown/March, and a pessimistic scenario where it is as high as in October, for each country. The infection curves for the three scenarios are well separated from each others for all three countries, indicating that each country might still further reduce its infection curves by decreasing its social interactions (by new measures or by changes in population behaviour) to values close to those during the first lockdown. In Luxembourg and Austria, the current level of social interactions seems to be sufficient to maintain a sub-linear growth of number of total cases (blue dashed curves in panels **A, B**, i.e. the number of daily cases decreases (panels **D, E** and **F**). However, for any of the three countries, an increase of the social interaction parameter to the levels of October would likely trigger a considerable rebound of the infection curve (red dotted curves in e.g. panels **D, E** and **F**). This will likely be associated with a rebound in the hospital (panels **G, H** and **I**) and ICU (panels **J, K** and **L**) occupation curves.

We see (panels **G, H, I** for hospitals, **G, H, I** for ICUs) that such a rebound to the social interaction levels of October could still lead to reach a peak in hospitals and ICUs occupations higher than those achieved in any of the past waves in the same country, and thus calls for the maximum caution by population and policy-makers alike.

It’s worth reminding that the number of dead is not only related to the height of the peak of daily cases, hospital or ICU occupations, but most importantly it is proportional to the area under these curves. This is obvious confronting panels **M, N** and **O** with the previous ones. We clearly see that keeping daily cases enough contained to avoid hospitals and ICU saturation, while necessary, does not prevent a steady growth of the cumulative number of dead. This number still grows significantly by spring in the simulations for all scenarios and countries, in particular it could still grow dramatically in the pessimistic scenario where we would go back to the social interaction levels of October.

### 4.2 Cross-validation of the model calibration and evaluation of parameter uncertainties

Complex epidemiological models often suffer from poor identifiability of their parameters [24]. This is associated with data quality as well as fitting methods. To account for this and provide consistent estimates, we employed two methodologies. First, we manually calibrated one parameters set for each wave and country, to fit the available time series data reasonably well. This is the parameter set we employ for the simulations of this work. Then, we cross-validated such choice with Bayesian inference, by means of Markov Chains Monte Carlo (MCMC) simulations. This elucidates parameters uncertainties, quantified by credible intervals, and their cross-correlation. A complete description of the Bayesian inference approach is reported in the Supplementary Material: Sec. S7 explains the methodology, and its detailed results are presented in Sec. S8 and figures therein.

Out of that analysis, three remarks are particularly relevant. First, that the uncertainties associated to each parameter are non negligible, and rather large for certain parameters (Fig. S2), possibly due to the large number of parameters. Second, that many parameter combinations are often better constrained than the corresponding single parameters (see e.g. Fig. S3). Third, that the manual fit is in good agreement with the Bayesian estimate (Fig. S2), but its set of parameters correspond to one possible choice and shouldn’t be considered fully exhaustive. These characteristics are common to any epidemiological model; here, we carefully investigated them, we incorporated domain knowledge from literature and we quantified their impact on our projections. Consequently, we recall that our estimates are not forecasts of the future, but rather likely assessments of potential scenarios.

### 4.3 Social interactions drive the dynamics of the epidemic and of *R*_*eff*_ (*t*)

The main parameter controlling the model behaviour is the social interaction *ρ* (*t*) (values in Tab. S3 for Luxembourg, Tab. S4 for Austria and Tab. S5 for Sweden), displayed in Fig. 3, left column. This modelling approach allows to fit available data, thus supporting the fact that, while any model is of course a simplification of the reality to its bare-bone crucial components, we could here confirm the well established fact that social interaction is a major driver necessary to understand the observed dynamics of the epidemic. In the model, this parameter represents both measures applied by policy-makers and behaviour of the population, thus underlining once more the importance of social behaviour in controlling the pandemic.

The values of the social interaction are inferred from model fit to available data. In turn, via Eq. (S6) derived analytically in Sec. S4, we can estimate variables and parameters the effective reproduction number *R*_*eff*_ (*t*), shown in Fig. 3, right column. The dynamics, i.e. the time-dependence, of this quantity is driven by only two factors: the social interaction parameter *ρ* (*t*) (causing step-wise changes in *R*_*eff*_ (*t*)), and the shrinking of the pool of susceptible *S* (*t*) (causing gradual changes in *R*_*eff*_ (*t*)). We see that so far the primary driver has indeed been the social interaction, in all three countries. The shrinking of *S* (*t*) starts to have a noticeable effect in Fall 2020 (*cf*. panels **B, D**, and **F** of Fig. 3), as for early times the population fraction (detected and undetected) who contracted the virus was still small compared to the entire population of the country. The effect of *S* (*t*) on *R*_*eff*_ (*t*) is more pronounced for Luxembourg than for Austria and Sweden, reflecting that a larger fraction of the total population has been infected in Luxembourg.

The reduction of the susceptible pool had a negligible effect during the first wave (even for countries not having imposed a lockdown as Sweden) and a mild effect at present. Nevertheless, the impact of shrinking *S* (*t*) is predicted to become visibly more relevant in the coming months as more people get infected, as can be seen from the stronger gradual bending of the curves in winter and spring 2021, especially for the pessimistic scenario in Fig. 3.

### 4.4 The epidemics in Luxembourg and Austria show similar dynamics, while Sweden exhibits very different dynamics

Our model could well fit Luxembourg and Austria, with parameter (probabilities and rates) changes only between subsequent waves, not during a wave. This is different for Sweden, which applied different policies than Luxembourg and Austria. We observed in Sec. 2.3 and Sec. S5.4 that in order to fit the data for Sweden with the same model structure, considering a different set of parameter values was not enough, instead we needed to introduce time-dependent probabilities of being detected *p*_1,*Swe*_ (Fig. S1, panel **B**) and being hospitalized *p*_2,*Swe*_ (Fig. S1, panel **D**), as detailed in Sec. S5.4.

From Fig. 2, we see that for Luxembourg and Austria the curves of new daily cases (panels **D** and **E**) follow the same dynamics as the curves of hospital and ICU occupations (panels **G, H, J** and **K**). This indeed supports our modelling choice, i.e. a model structure leading to the same dynamics for these compartments, simply scaled and delayed. On the other hand, for Sweden we see that during the first wave the dynamics of new daily cases (panel **F**) was very different than that of hospital and ICU occupation (panels **I** and **L**, having similar dynamics between each other).

These different dynamics between detected cases and hospitalisations in Sweden does not reflect a rise in infections, but rather a major change in testing strategy in Sweden which changed dramatically during May 2020, leading to an increase in detected cases. As observed in [33], the peak recorded towards the end of June reflects an extension of the testing strategy to include everyone with the symptoms of COVID-19, as well as testing to take place in cases of contact tracing. Furthermore, the number of daily tests performed increased significantly in late May and the following weeks, due to a decision by the government [28]. In order to incorporate this, we had to make the parameter *p*_1_ time-dependent.

Moreover, for our model to fit the data of Sweden, we also had to make the probability of being hospitalized when detected positive *p*_2_ time-dependent. In fact, for Sweden the ratio of people entering hospitals divided by new daily detected cases two days earlier (Fig. S1, panels **C, D**) seemed to be considerably above 50% (close to 100% at times) during March and April, meaning that in the early weeks/months of the epidemic in Sweden mainly severe cases requiring hospital treatment were detected. This clearly changed in May and June, likely due to the change in testing strategy [28].

### 4.5 First and subsequent waves exhibit differences in hospitalization probability

For Luxembourg, Austria and Sweden, the set of fitting parameters for the first wave does not allow to fit the second one, despite the model structure being conserved.

With the manually calibrated parameter sets, we test if such a change could be explained primarily by lowered values of *p*_2_, and observe that indeed this could be the case. For Luxembourg (*cf*. Tab. S6 and Fig. 4), it decreased from 15.3% in 1st wave to 8.0% in 2nd wave to 4.2% in 3rd wave. For Austria, it was decreased from 16.2% in the 1st wave to 6.5% in the 2nd wave. In Sweden, we assumed for it a time-dependent (piecewise linear) behavior, decreasing from 90% in March to 10% from June onward. These changes where also supported by similar changes in the maxima of the Bayesian estimates for 3^rd^ versus 2nd wave in Luxembourg and 2nd versus 1st wave in Austria. We hypothesise that this considerable decrease of the probability of being hospitalized once detected might reflect the combination of considerably increased testing efficiency and better prognosis linked to younger patient age.

### 4.6 Vaccination might lead to herd immunity in 2021, the timing will depend on vaccination strategy and social interaction

After extending the model with a vaccinated compartment, we quantified how much time is plausibly necessary until enough people are vaccinated for herd immunity.

Our model contains compartments for the undetected cases, the parameters of which are estimated based on prevalence data. Luxembourg and Austria display (*cf*. Fig. 2) a similar fractions of undetected, which are approximately 1.5 to 2 times bigger than the number of detected, depending on the date, while Sweden has a larger number of undetected cases, approximately 4 times the detected cases, accumulated mostly during the first wave. This is important to assess the true infected population fraction, towards herd immunity. Until 15th December in our model, in Luxembourg about 18.3% (7.2% detected and 11.1% undetected) of the population had SARS-COV-2; in Austria 9% had the virus (3.7% detected and 5.3% undetected), while for Sweden 14.5% had the virus (3.5% detected and 11% undetected). Despite the different policies, Sweden is actually not only far from herd immunity, but also further than e.g. Luxembourg.

We observed in Fig. 5 that the difference between the detected cases curves of the two considered social interaction scenarios is larger than the difference between the different vaccination strategies, or of comparable size at best. This indicates that during the vaccination process, reduced social interaction is still going to be crucial in order to keep under control the number of cases, and thus not to saturate hospitals and ICUs and to reduce the number of deaths. Therefore, even during the initial months of vaccination, social behaviour should not be relaxed too much.

Considering herd immunity by vaccination only, we observe in the first row of Fig. 6 that for Luxembourg the strategy under which everyone get vaccinated within 6 months allows to reach herd immunity by April, while in Austria and in Sweden this strategy is not enough to get it in April. Slower strategies will not lead to herd immunity in April unless values of the social interaction parameter are at least as high as in October. However, such values are undesirable due to their consequences.

We further found that, to reach herd immunity by mid April primarily by vaccination (starting on January 1st), Luxembourg will need approximately a bit less than 3000 vaccinations/day, Austria and Sweden approximately 45000. Considering average (w.r.t. March-December 2020) social interaction, Luxembourg needs at least 2500 vaccinations/day, Austria and Sweden 45000. If social interaction would go back to the (high) levels of October, at least 2000 vaccinations/day would anyway be needed for Luxembourg and 30000 for Austria and Sweden. Nevertheless, at a more fine grained look we see that the combination of social interaction and vaccination leads to subtle differences between these last two countries. If the social interaction parameter would be 0.4, Austria would need approximately 13000 vaccinations/day, thus more than Sweden needing 12000, in order to reach herd immunity by mid April. Conversely, for a social interaction parameter of 0.5 Austria would need approximately 38000 while Sweden a smaller number, namely 32000.

Moreover, in the second row of Fig. 6 we see that for Luxembourg the strategy of vaccinating everyone in 6 months, alone, would provide herd immunity by April, while for Austria and Sweden it will hardly be enough. The other two strategies will not suffice for herd immunity by April, except in the case where the social interaction parameter would be as high as in October, which would on the other hand lead to an undesirably large wave of infections with its consequences in terms of dead and so on.

We also find that, if vaccinating all country’s population within 1 year at a constant rate, to reach herd immunity by vaccination alone Luxembourg would take until July, Austria until August and Sweden early September. With an average value of social interaction together with this vaccination strategy, herd immunity could be reached in June in Luxembourg and July in Austria and Sweden. If social interaction would go back to the levels of October 2020, it would anyway take until April for all three countries to reach herd immunity, but with a much greater burden for the health care system and in term of lives.

Overall, while vaccination will surely be helpful in protecting the vulnerable, limiting social interactions will still play a major role until the vaccination effects become relevant in the pursuit of herd immunity during the course of 2021.

### 4.7 Limitations of this work and potential future extensions

In this study we investigated three countries: Luxembourg, Austria with a similar dynamics, and Sweden having a different dynamics. While this already showcase a range of interesting situations, we could extend the model to other countries, in particular those with very different properties.

In our cross-validation of the model calibration, we assumed flat prior probability distributions over a parameter space as wide as reasonably possible, *cf. Sec. S7.2*, in order to investigate the constraints on the parameters provided by the available data. A different set of priors could have been considered, e.g. Gaussian priors centered around the manually fit parameter values. Nevertheless, we didn’t perform this as we didn’t want to introduce bias in the Bayesian parameter estimate meant to provide crossvalidation of the manually calibrated parameter set. The likelihood function we built from the sum of square residuals attribute the same weight to all time-series data. While this choice is the simplest, it is not unique and more complex weighting approaches could have been considered.

Moreover, we should stress here that our model projections, while potentially accurate in the short term, for longer time-periods are to be intended as a way to compare potential scenarios and gain insight. In fact, SEIR-based model simulations are known [11] to be sensitive to parameter values which can either change in time or be affected by large uncertainties in their estimate.

While SEIR-like models were already investigated before the pandemic for their interesting mathematical properties e.g. in [34], alternative approaches to differential equation based SEIR-like models have been widely employed, as e.g. the age-stratified discrete compartment model for Switzerland including the different phases of disease progression for infected individuals [35]. Models based on ordinary differential equations, as the SEIR and most of its extensions as ours, are deterministic in nature. Thus, they do not accurately describe the stochastic aspects of epidemics, e.g. super-spreading events. These are better captured by agent-based models [36], which have widely been used in modelling COVID-19 epidemic. Deviations from mean-field effects are expected to play an important role during periods of low numbers of infections, where e.g. one or few super-spreading events can make the difference between starting or not a wave of infections. However, with high case numbers we expect a deterministic description to be accurate enough, especially when it comes to the progression of infected individuals into more severe stages as hospitalisation, ICU and death.

Each country is here modelled as a closed system, which might be a limitation particularly for Luxembourg, due to the small size of the country and the high cross-border mobility. The data employed for Luxembourg only include cases that are detected and resident in Luxembourg. For considerations about cross-border workers in Luxembourg and interplay with economical features, refer to [10].

In the present model, people who recover from the infection are considered to be immune and thus cannot be re-infected. While it seems that individuals who have been infected with COVID-19 tend to develop some level of immunity to the disease for a certain time [18], the duration of this immunity is not yet clear. Cases of re-infection of the same individual have been reported [37]. Re-infection by coronaviruses in general was found to occur frequently [38], but the same study stresses that there is limited evidence of reinfection by SARS-CoV-2. Persistence and decay of antibodies to SARS-Cov-2 was investigate in [39], and [31] found that asymptomatic individuals had a weaker immune response to SARS-CoV-2 infection. The assumption that appearance of SARS-CoV-2 antibodies provides protection from reinfection, implicit in SEIR-like models, is questioned in [40]. Nevertheless, overall re-infection by SARS-Cov-2 has only been conclusively documented in a very limited number of cases, and it is unclear to date whether this is a rare phenomenon or may prove to become a common occurrence [17]. We have here considered reinfection to play a negligible role for the purposes of this work. Nevertheless, reinfections might be relevant on the time scale of years.

Furthermore, the model could also be extended to include age-distribution, particularly relevant when focusing on ICUs and dead. Moreover, it has been shown in a model including age-structured community with mixing rates fitted to social activity [32] that the disease-induced herd immunity level can be lower than what we considered here, in particular around 43% for *R*_0_ ≈ 2.5. This would frame our estimates as a worst-case. On the other hand, vaccine efficacy and duration of protection are key issues, which we do not include here (thus assuming a vaccine with 100% efficacy that gives life-long protection). Both would eventually contribute to make herd immunity more challenging to reach [21], and little is known on the duration of protection, even if in principle vaccination could be repeated regularly.

Finally, we remark that our estimates of herd immunity being achieved by vaccination assume that the current vaccines successfully prevent the vaccinated individuals from spreading the virus, on top of protecting them from developing the COVID-19 disease. Up to date, this is not yet fully assessed, but it is a main goal of a vaccination campaign and it is therefore of the out-most importance to investigate it.

## 5 Conclusions

An extended SEIR model was developed in order to incorporate social interaction, undetected cases, progression through hospital, ICU, recovery, death and vaccination. It was used to provide projections, gain insights on the different wave dynamics comparing within and between countries, and investigating the interplay of social interaction and vaccination in the pursuit of herd immunity by vaccination.

We fit the model independently to Sweden, Luxembourg and Austria. The epidemics in Luxembourg and Austria show similar dynamics, with the model fitting both with appropriate changes in parameter values. The epidemic in Sweden exhibits instead a very different dynamics. In order to fit Sweden with the same model structure, time-dependent probabilities of being detected and being hospitalized are necessary. This highlight, first, a major change in test strategy occurring in Sweden in May 2020, and second, that the majority of the detected cases in the early months in Sweden were people needing hospital care.

While several parameters can be used across waves and countries, some parameter values need adjustments between subsequent waves in the same country. In particular, we remark that the probability of being hospitalized if detected might have become lower with subsequent waves, in all three countries considered here, possibly due to considerably improved testing strategies and improved prognosis of patients due to their younger age.

Our model provides projections for potential future scenarios. We observe that an increase of the social interaction parameter to the levels of October (e.g. due to relaxation of measures or major relaxation in people’s behaviour) could still result for all three countries in dramatic increase in the number of detected cases, hospital and ICU occupation and dead.

Finally, we employed our model to investigate the interplay between social interaction and vaccination, in view of obtaining herd immunity. While vaccination will surely help to protect the most vulnerable, during the early months of 2021 social interaction will still contribute more or as much as vaccination to mitigate the infection curve. The estimated number of undetected cases, accounted for in the model, turns out to be larger for Sweden than for Austria and Luxembourg, while Luxembourg is the country with the highest fraction of people (detected and undetected) who have been infected so far. Nevertheless, all three countries are far from herd immunity on 15th December 2020, having an estimated 14.5 % of the population having been infected in Sweden, 9% in Austria and 18.3% in Luxembourg. Herd immunity is within reach for 2021, but it will unlikely be reached in the early months of the year, and it might take until the end of the year or even until 2022 to reach it, depending on how fast countries can vaccinate their population. Meanwhile, social interaction will still be a major driver of the pandemic.

On the one hand, the challenge over the coming year will be for governments and populations to use COVID-19 vaccines efficiently to create herd immunity to protect all. On the other hand, it will be to keep social interaction limited, as this will still remain crucial during 2021 before vaccination might be able to provide us with herd immunity.

### Ethics

The application of anonymized data for the purpose of epidemic modelling has been endorsed by the accessed databases.

### Code accessibility

The code implementing the model and analysis will be released upon journal publication.

## Data Availability

All data employed are publicly available at the links indicated in the manuscript.

## Authors’ contributions

FK, SM and DP designed the study. FK, SM and DP developed the model. FK and SM implemented the model and performed the simulations. FK, SM, DP, AH, AA, AFDH, LM, AS, JG, CL analyzed and interpreted the results. SM, AH, AS, JG, CL supervised and coordinated the project. FK, SM, JG, AS, CL wrote the first draft. All authors contributed to the final draft. All authors gave their final approval for publication.

## Acknowledgments

The authors thank the Research Luxembourg - COVID-19 Taskforce for mutual collaborations. We are also grateful to Matías Nicolás Bossa for useful discussion.

## Supporting material: model and methods

In this supporting material, we illustrate the model we developed, the data we considered from Luxembourg, Austria and Sweden, and the procedures we employed to provide an estimate of the parameters and their uncertainties.

### S1 Mathematical model

In this study, we developed a mathematical model of the transmission of COVID-19 within a population, building upon the standard SEIR model [22]. Our extension of the SEIR model is described by Fig. 1 in the main text. First, we introduce in the model a separation between detected and undetected cases of COVID-19, to represent the fact that a non-negligible fraction of infected people is not detected [23], e.g. for lack of testing or because asymptomatic. Next, we include compartments to model the progression of the disease in individuals who have been infected, in particular the progression to more severe states, requiring hospitalisation, intensive care (ICU) or leading to death, and finally compartments for recovered people.

The total population *N* of the modelled country at time *t* is divided into 16 compartments. The variables are summarized in Tab. S1 and represent the number of individuals in different stages, normalized by the total population of the modelled country. Susceptible (*S*) and exposed (*E*) compartments are in common with the standard SEIR model. An exposed person might be detected (via a PCR test), thus entering the “infectious and detected” (*II*) compartment, and subsequently the “quarantined” (*Q*) compartment, from where the person cannot infect others any further. At this point, a person might (or not) become “hospitalized” (*H*), and then either simply “require longer hospitalization” (*Hl*) or require “intensive care treatment” (*ICU*). If the health improves, the individual can return to “regular hospital treatment after ICU” (*AICU*). At any of these hospitalization stages, the person can “die in hospitals or ICUs” (*D*_*I,hos*_), or “recover from hospital or ICUs” (*R*_*II*_). People who are either recovered or dead are the equivalent of the removed compartment of a standard SEIR model, and thus do not get reinfected a second time in the model. Most detected people are not hospitalized, so they recover at home (*NI*), or alternatively they can die at home (*D*_*I,hom*_). Finally, the fraction of people who got infected but do not get detected are represented by the infectious and undetected compartment (*A*). Being unaware of their state, they can continue to spread the infection without constraints. These people spend some time recovering outside hospitals (*N*_*II*_), and eventually they end up either recovered undetected (*R*_*A*_), or dead undetected (*D*_*A*_).

We assume conservation of the total population *N* (people can die, thus entering the dedicated compartments, but there is no removal of individuals from the system, and we neglect birth), such that for each country:

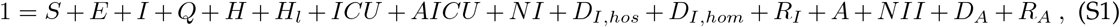

where the variables have been normalized by *N* to represent the fraction of the total population being in the corresponding state.

We initialise every simulation with the initial conditions detailed in Tab. S1. For any simulation in this work, to numerically solve the set of ordinary differential equations which implements the model we employ the python (version 3.6.1) solver odeint from the package scipy (version 1.5.2), which uses the LSODA method for numerical integration. This is an Adams/BDF method with automatic stiffness detection and switching, which we have chosen for both speed and precision. It is in turn a wrapper to the Fortran solver from ODEPACK [41]. It switches automatically between the non-stiff Adams method and the stiff BDF method. The method was originally detailed in [42].

The dynamics of our model is described by the following system of ordinary differential equations:

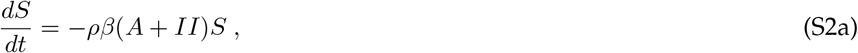

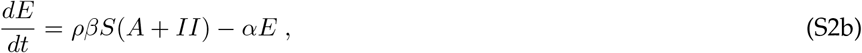

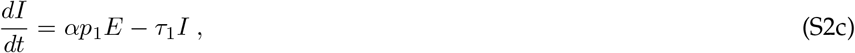

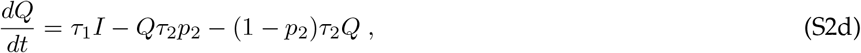

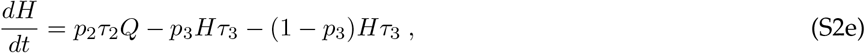

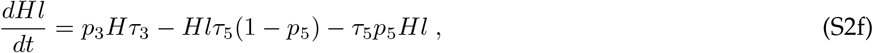

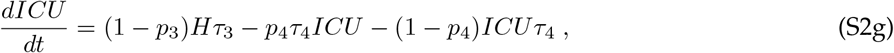

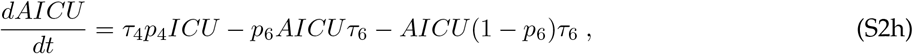

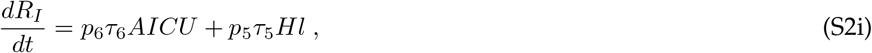

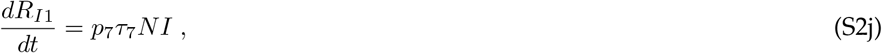

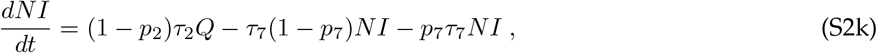

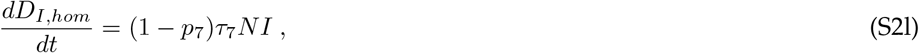

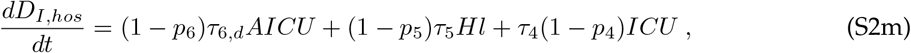

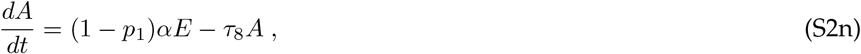

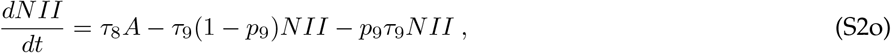

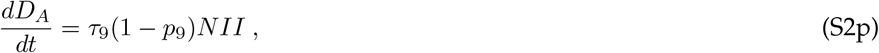

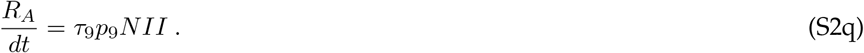

### S2 Model parameters

The parameters of the model and their interpretation are detailed in Tab. S2. The parameters can be divided in three categories, based on what type of physical quantity they describe: rates, probabilities and social interaction.

When individuals can move from one compartment to one of two different compartments, the probability *p*_*i*_ controls which fraction of people will go to one compartment and which to the other. These parameters represent the probability of going into a compartment from the previous, and thus the probability of going to the alternative compartment is given by 1 − *p*_*i*_. Their value is bounded to be between 0 and 1. The parameters *p*_*i*_ are indexed with *i* = 1, …, 9.

The parameters *α, β* and *τ*_*i*_ represent rates. The parameters *τ*_*i*_ are indexed with *i* = 1, …, 9. Each rate represents the inverse of the average time that takes for an individual to move from one compartment to the next. Of particular relevance are the rates *α* and *β*, which have the same meaning as the corresponding parameters of a standard SEIR model: *α* represents the inverse of the mean incubation period of the disease, and *β* represents the average contact rate.

**Table S1:**
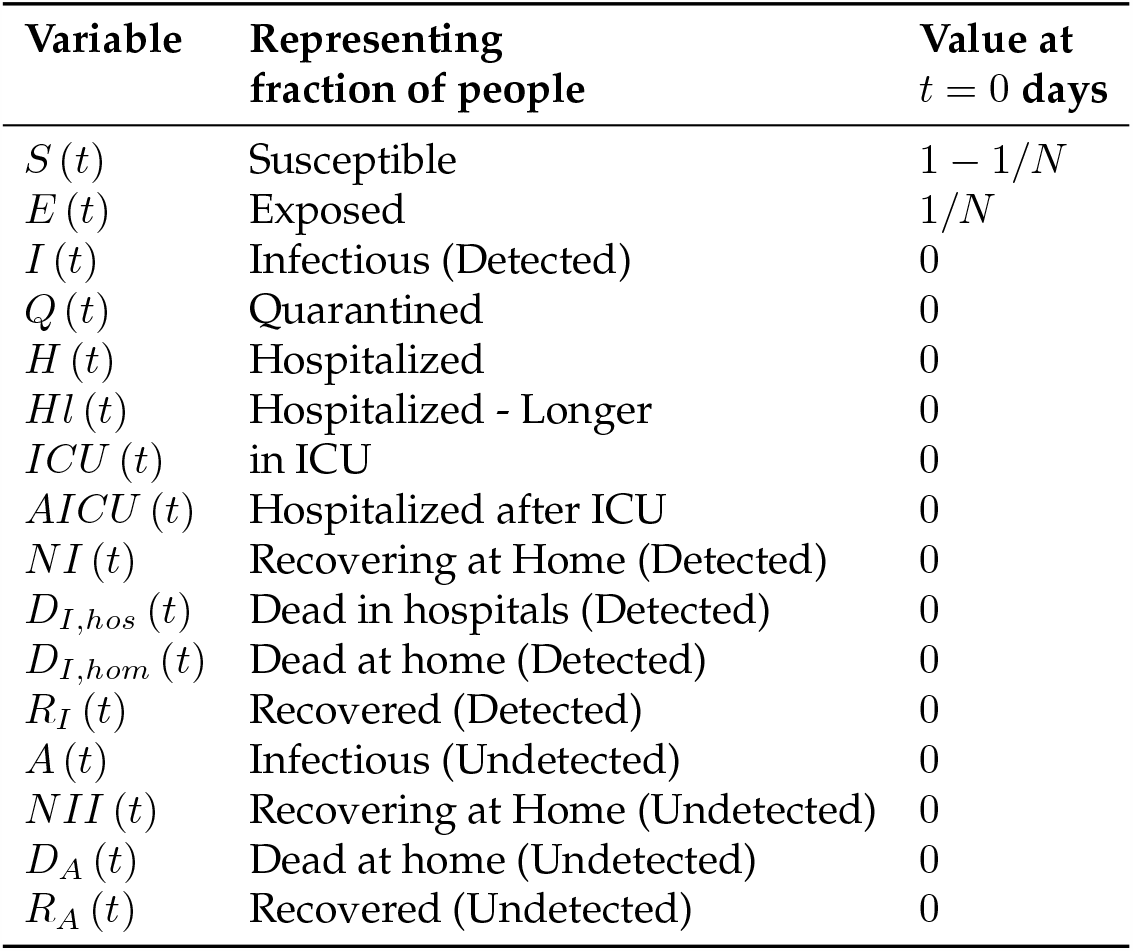
Variables employed in the model and their initial conditions. The 16 variables represent the fraction of population of a country being in each of the compartments. The initial conditions are the standard ones usually employed for SEIR-like models, with all the population susceptible except one person already exposed to the virus. They are the same for every simulation and country, except for the country’s population being *N* = 623180 individuals for Luxembourg, *N* = 8901064 for Austria and *N* = 10230000 for Sweden.

The average contact rate *β* is then multiplied by the additional parameter *ρ*, in order to model any measure or change in people’s behaviour that can lead to a change (decrease or increase) in the average contact rate. Thus, *β* is a constant and it represents the “natural” average contact rate, while *ρ* (*t*) *β* represents an effective average contact rate which considers the measures or social behaviours in place. We assume *ρ*_0_ = 1 for any country, before any measure was implemented at the beginning of the pandemic in February 2020. Afterwards, we assume *ρ* = *ρ* (*t*) to be a piece-wise constant function of time, where each value is indicated by *ρ*_*n*_, with *n* = 0, …, 13 for Luxembourg and Sweden and *n* = 0, …, 16 for Austria. We assume that changes in *ρ* occur whenever a major new measure is implemented or lifted by the authorities of a country, or in case of a major happening (e.g. schools starting in September in Luxembourg). The dates employed for every country, and which measures where taken/lifted on that date, are summarized in Tab. S3 for Luxembourg, Tab. S4 for Austria and Tab. S5 for Sweden.

### S3 Incorporating vaccination in the model

To include in the model vaccination, which we investigate in Sec. 3.4, we add one additional compartment *V* (*t*) representing the fraction of vaccinated population. We assume that a fixed number of people will be vaccinated every day. Thus, the equation for the number of people vaccinated reads:

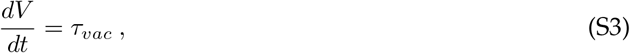

with *τ*_*vac*_ being the parameter representing how many people will be vaccinated per day. Correspondingly, the equation for the susceptible compartment becomes:

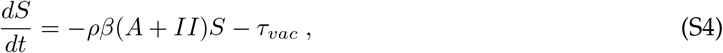

since we assume that only people who did not already naturally develop antibodies by being infected will be vaccinated.

**Table S2:**
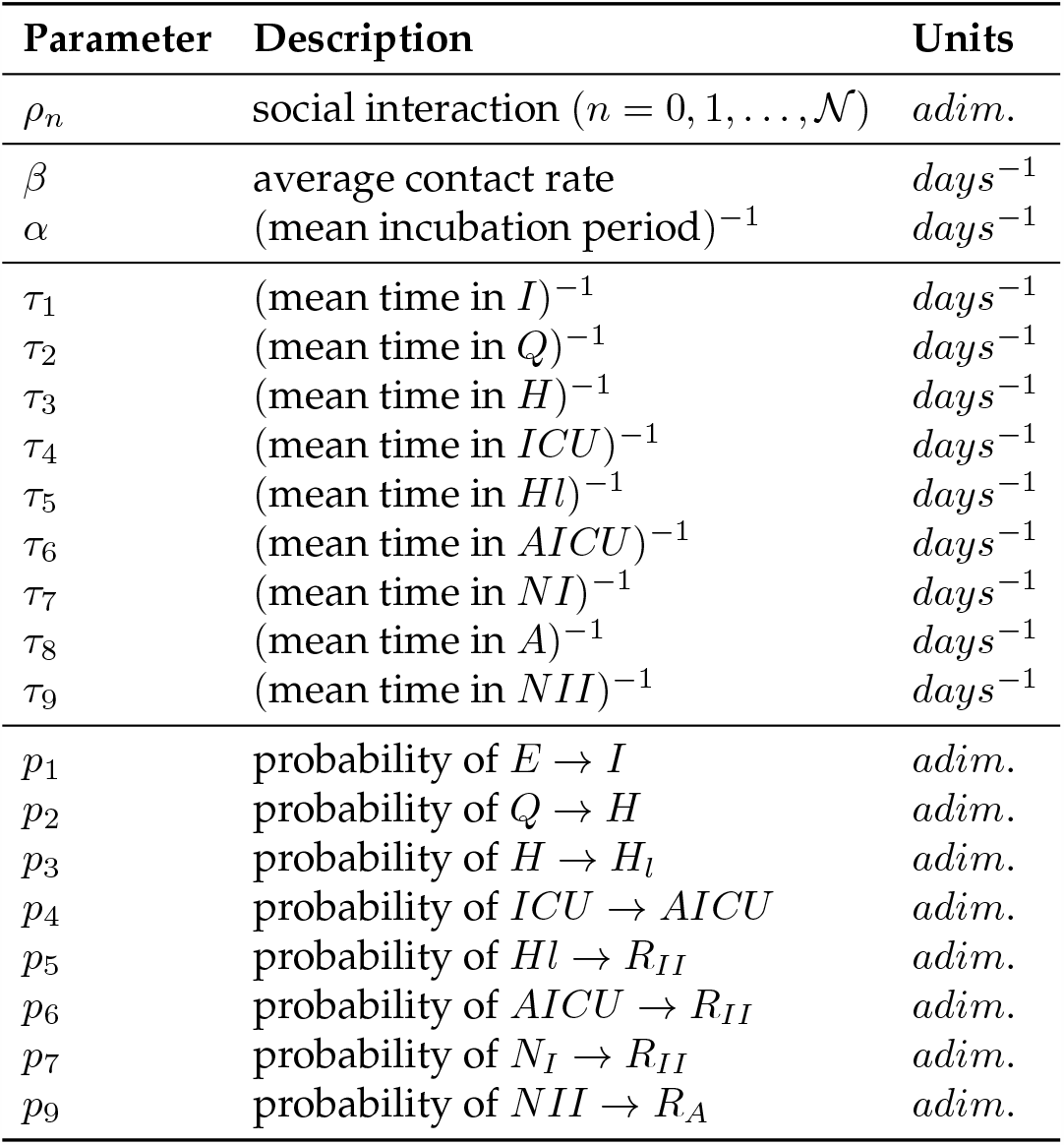
Parameters of the model. The parameters of the model with their description and unit of measure. For each country, the following assumptions have been made: *τ*_8_ = (1*/τ*_1_ + 1*/τ*_2_)^−1^, *p*_9_ = *p*_7_, *τ*_9_ = *τ*_7_. The same assumption is made for each wave. This is motivated by the fact that these parameters are relative to the undetected branch, and thus cannot be inferred from data. Thus, we assumed that undetected cases would evolve with the same parameter values as detected cases not entering hospitals/ICUs. 𝒩 represents the number of times that the social interaction parameter changes: 𝒩 = 13 for Luxembourg and Sweden and 𝒩 = 16 for Austria. The values of the manual fit of all the rates and probabilities for each country and wave are summarized in Tab. S6. The values and dates of change for the social interaction parameter *ρ*_*n*_ are reported in Tab. S3 for Luxembourg, in Tab. S4 for Austria and in Tab. S5 for Sweden.

In order to simulate three potential vaccination strategies, i.e. 3 speeds at which countries might manage to perform vaccination, we fix the parameter *τ*_*vac*_ to three values that, if all the population would still be susceptible, would lead to vaccinating all the population of a country within respectively 6 months, 1 year and 1.5 years. These are typical timescales potentially envisaged by different countries. Thus, we consider the three values *τ*_*vac*_ = 1*/* (365*/*2) *days*^−1^, *τ*_*vac*_ = 1*/*365 *days*^−1^ and *τ*_*vac*_ = 1*/* (1.5 · 365) *days*^−1^. It should be recalled that model variables are normalized by the total country population *N*, so they sum up to 1. These three values of *τ*_*vac*_ correspond, respectively, to perform approximately 3415, 1707 and 1138 vaccinations/day in Luxembourg, 48773, 24386 and 16258 vaccinations/day in Austria and finally 56055, 28027 and 18685 vaccinations/day in Sweden.

When numerically integrating the system of ordinary differential equations from Eq. (S2), that constitutes the model, with the addition of vaccination (Eq. (S3) and Eq. (S4)), we also need to add an additional constraint to prevent the variable *S* (*t*) to decrease below 0, which would happen otherwise due to Eq. (S4). We employed everywhere else in this work the odeint routine from the scipy python library to perform numerical integration. Nevertheless, that routine does not allow to dynamically interrupt the integration when a certain condition is met (*S* = 0) to prevent it from decreasing further by setting *τ*_*vac*_ = 0 from that time on. Thus, in all the simulations involving vaccination we instead perform numerical integration by means of the forward Euler method. In fact, in order to compute the integral of the ODE system for a given time this method does not need to evaluate the system at subsequent times, and thus allows changing a parameter at a time-step based on the outcome of the integration at the previous time-step. We employed a time-step of integration *dt* = 0.01 *days* and verified beforehand that further decreasing it would not lead to any significant change in the integration result, while significantly increasing computation time.

### S4 Analytical derivation of the effective reproduction number *R*_*eff*_ (*t*) for the model

To obtain *R*_*eff*_ (*t*) from the model we used the next generation matrix method [43, 44, 45]. From the next generation matrix, we find its eigenvalues *R*_0_ and 0. In our case, *R*_0_ is given by:

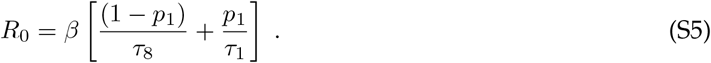

From *R*_0_, we can deduce *R*_*eff*_ (*t*), given by

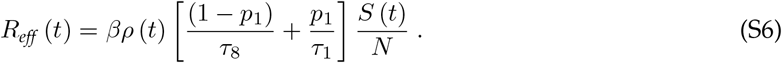

By substituting the values of the parameters for each country and scenario, we obtain the *R*_*eff*_ (*t*) curves depicted in Fig. 3, panels **B, D, F**.

### S5 Data and analyzed countries

We employ our model to describe the progression of the pandemic in three different countries: Luxembourg, Austria and Sweden. The model structure described above is maintained unchanged across the three countries. In order to make the model describe appropriately each of the countries, we instead change the value of the total country population *N*, and the parameter values used in the model so that the model fits the data of the corresponding country. We consider data of total detected cases, hospitalized people, people in ICUs and dead people. We gathered these data from^3^ and integrated them for each country with further data from public repositories listed below. Moreover, for each country we allow the social interaction parameter *ρ* to change when major changes in measures took place. We list below also the sources of information for the changes in measures.

#### S5.1 Luxembourg

For Luxembourg, we obtained the dates at which major changes in policy took place in Luxembourg from ^4^, and summarize them in Tab. S3. The time-series data of total cases, hospitalised people, people in ICU and dead people for Luxembourg are publicly available on^5^. They are reported in Fig. 2, along with their 7-days moving average. This moving average is necessary, not only to smooth noise due to detection and to the nature of the data itself, but mainly to remove the effect of a considerable under-testing on weekends, and in particular on Sundays, which has been observed every week in the reported daily detected cases and which is confirmed by the considerably lower number of tests performed (positive or negative) over these weekend days. We point out that we employ a moving average which, for each day, computes the average between the values from 3 days before to three days after, thus not introducing any shift of the features (like the peaks) of the time-series.

According to the public data mentioned above, the first positive case was detected in Luxembourg on February 29, 2020. Considering the lag between being susceptible, being exposed and being detected, we initialize the model at its initial conditions reported in Tab. S1 with time *t* = 0 on February 24.

Data about the prevalence in Luxembourg’s population were obtained from the Con-Vince study [46].

**Table S3:** Changes in the piece-wise constant social interaction parameter *ρ* (*t*) of the model for Luxembourg.

| $\rho_n$ | Starting Date | Measure or change in social activities | Manual Fit value | Which Wave |
| --- | --- | --- | --- | --- |
| $\rho_0$ | — | No restrictions | 1 | — |
| $\rho_1$ | 16.03 | Lockdown | 0.4902 | 1st |
| $\rho_2$ | 23.03 | Measures fully effective | 0.1984 | 1st |
| $\rho_3$ | 20.04 | Opening construction sector | 0.1918 | 1st |
| $\rho_4$ | 04.05 | Opening high schools | 0.2507 | 1st |
| $\rho_5$ | 10.06 | Relaxation of measures | 0.5298 | 2nd |
| $\rho_6$ | 29.06 | End of classes splitting | 0.4804 | 2nd |
| $\rho_7$ | 13.07 | School holidays | 0.3101 | 2nd |
| $\rho_8$ | 31.07 | Start holidays | 0.2203 | 2nd |
| $\rho_9$ | 12.08 | Change mobility (Google mobility data) | 0.3457 | 2nd |
| $\rho_{10}$ | 23.08 | End holidays of construction sector | 0.3351 | 2nd |
| $\rho_{11}$ | 14.09 | Opening schools | 0.4333 | 3rd |
| $\rho_{12}$ | 01.11 | Restrictions, including curfew (11 p.m.) | 0.341 | 3rd |
| $\rho_{13}$ | 26.11 | Stricter measures, HORESCA closure | 0.39 | 3rd |

#### S5.2 Austria

We obtained the dates at which major changes in policy took place in Austria from ^6^, and we summarize the dates and the policy changes in Tab. S4. The time-series data of total cases, hospitalised people, people in ICU and dead people for Austria come from^7^. They are reported in Fig. 2, where we also report their 7-days moving average.

**Table S4:**
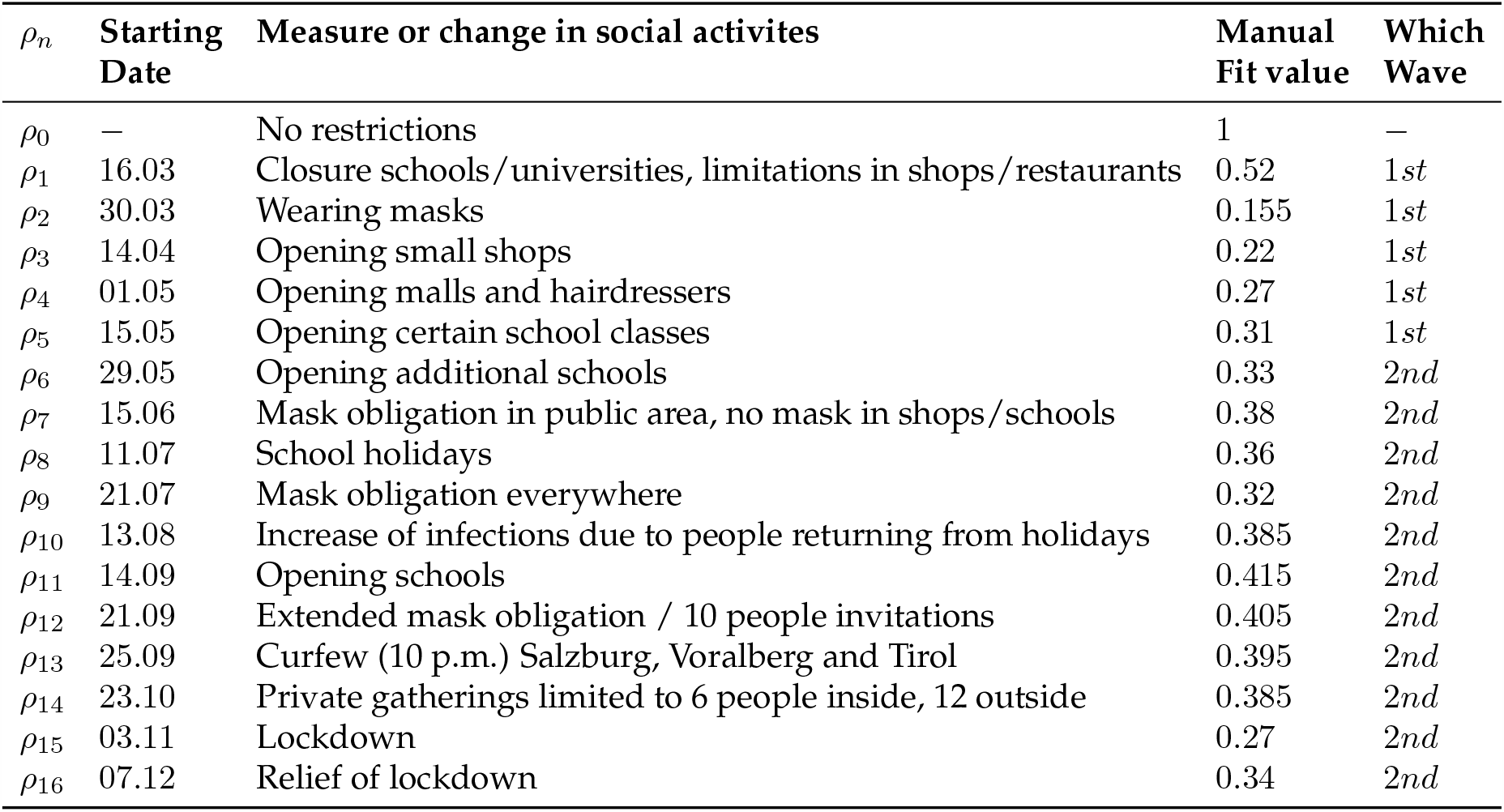
Changes in the piece-wise constant social interaction parameter *ρ* (*t*) of the model for Austria.

| $\rho_n$ | Starting Date | Measure or change in social activities | Manual Fit value | Which Wave |
| --- | --- | --- | --- | --- |
| $\rho_0$ | — | No restrictions | 1 | — |
| $\rho_1$ | 16.03 | Closure schools/universities, limitations in shops/restaurants | 0.52 | 1st |
| $\rho_2$ | 30.03 | Wearing masks | 0.155 | 1st |
| $\rho_3$ | 14.04 | Opening small shops | 0.22 | 1st |
| $\rho_4$ | 01.05 | Opening malls and hairdressers | 0.27 | 1st |
| $\rho_5$ | 15.05 | Opening certain school classes | 0.31 | 1st |
| $\rho_6$ | 29.05 | Opening additional schools | 0.33 | 2nd |
| $\rho_7$ | 15.06 | Mask obligation in public area, no mask in shops/schools | 0.38 | 2nd |
| $\rho_8$ | 11.07 | School holidays | 0.36 | 2nd |
| $\rho_9$ | 21.07 | Mask obligation everywhere | 0.32 | 2nd |
| $\rho_{10}$ | 13.08 | Increase of infections due to people returning from holidays | 0.385 | 2nd |
| $\rho_{11}$ | 14.09 | Opening schools | 0.415 | 2nd |
| $\rho_{12}$ | 21.09 | Extended mask obligation / 10 people invitations | 0.405 | 2nd |
| $\rho_{13}$ | 25.09 | Curfew (10 p.m.) Salzburg, Voralberg and Tirol | 0.395 | 2nd |
| $\rho_{14}$ | 23.10 | Private gatherings limited to 6 people inside, 12 outside | 0.385 | 2nd |
| $\rho_{15}$ | 03.11 | Lockdown | 0.27 | 2nd |
| $\rho_{16}$ | 07.12 | Relief of lockdown | 0.34 | 2nd |

According to the data mentioned above, the first positive case was detected in Austria on February 26, 2020. Considering the lag between being susceptible, being exposed and being detected, we initialize the model at its initial conditions reported in Tab. S1 on February 21. Similar starting dates for modelling the epidemic in Austria have been used elsewhere, with [14] starting their modelling for Austria on February 22, see Tab. I in [14].

Data about the prevalence in Austria were obtained from^8^.

#### S5.3 Sweden

We obtained the dates at which major changes in policy took place in Sweden from [28] until August and from ^9^ later on, and we summarize them in Tab. S5. The time-series data of total cases, hospitalised people, people in ICU and dead people for Sweden are available on^10^. They are displayed in Fig. 2, where we also report their 7-days moving average.

Swedish policy has been analysed e.g. in [33, 47, 48]. While the very first case detected was on 31st January, no other case was detected until February 26. It is likely that the first case was isolated and did not infect others, otherwise most likely a second case would have been detected earlier; in addition, the first case was reported from a woman who had traveled to Wuhan and was isolated upon detection. On the other hand, following the second detected case on February 26, 9 more cases were detected in the next 2 days. To account for potential further lag in the uncertain starting time of the epidemic in Sweden, we initialise the model at its initial conditions reported in Tab. S1 on February 16. Similar starting dates for modelling the epidemic in Sweden have been used elsewhere, with [14] starting their modelling on February 18, see Tab. I in [14].

**Table S5:**
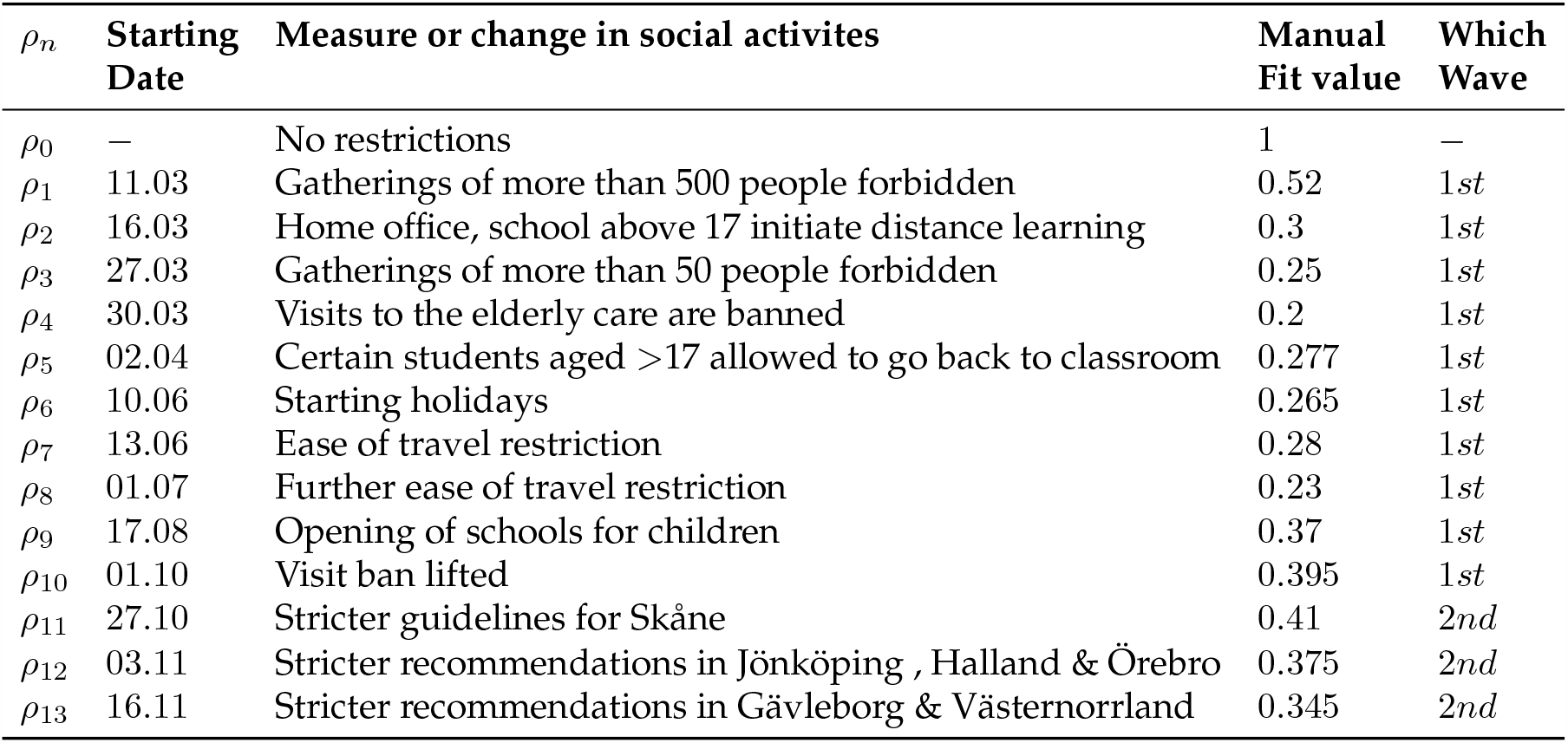
Changes in the piece-wise constant social interaction parameter *ρ* (*t*) of the model for Sweden.

#### S5.4 To fit Sweden, the model needs time-dependent probabilities of detection and hospitalisation, unlike for Luxembourg or Austria

We extend our model to Sweden and compare its simulations and the model-building process, to gain insight on a country which applied different policies than Luxembourg and Austria (which adopted similar policies). To fit Swedish data with the same model structure as the other countries, it was not sufficient to consider a different set of parameter values. Instead, it was necessary to introduce time-dependent probabilities of being detected *p*_1Swe_ (*t*) and of being hospitalized *p*_2Swe_ (*t*), within an epidemic wave.

The parameter *p*_1_ corresponds to the probability of an infected individual being detected. To obtain an estimate of this parameter for Sweden, we considered prevalence data estimated weekly through antibody test for 8 consecutive weeks between mid April and mid June 2020, obtained from [25, 26] and depicted in Fig. S1 panel **A**. The prevalence went from about 4% to 6%, growing over the course of these two months, likely resulting from more infections. Due to the large error bars, multiple functional forms could be considered a reasonable fit, so we chose the simplest linear fit.

**Fig. S1.**
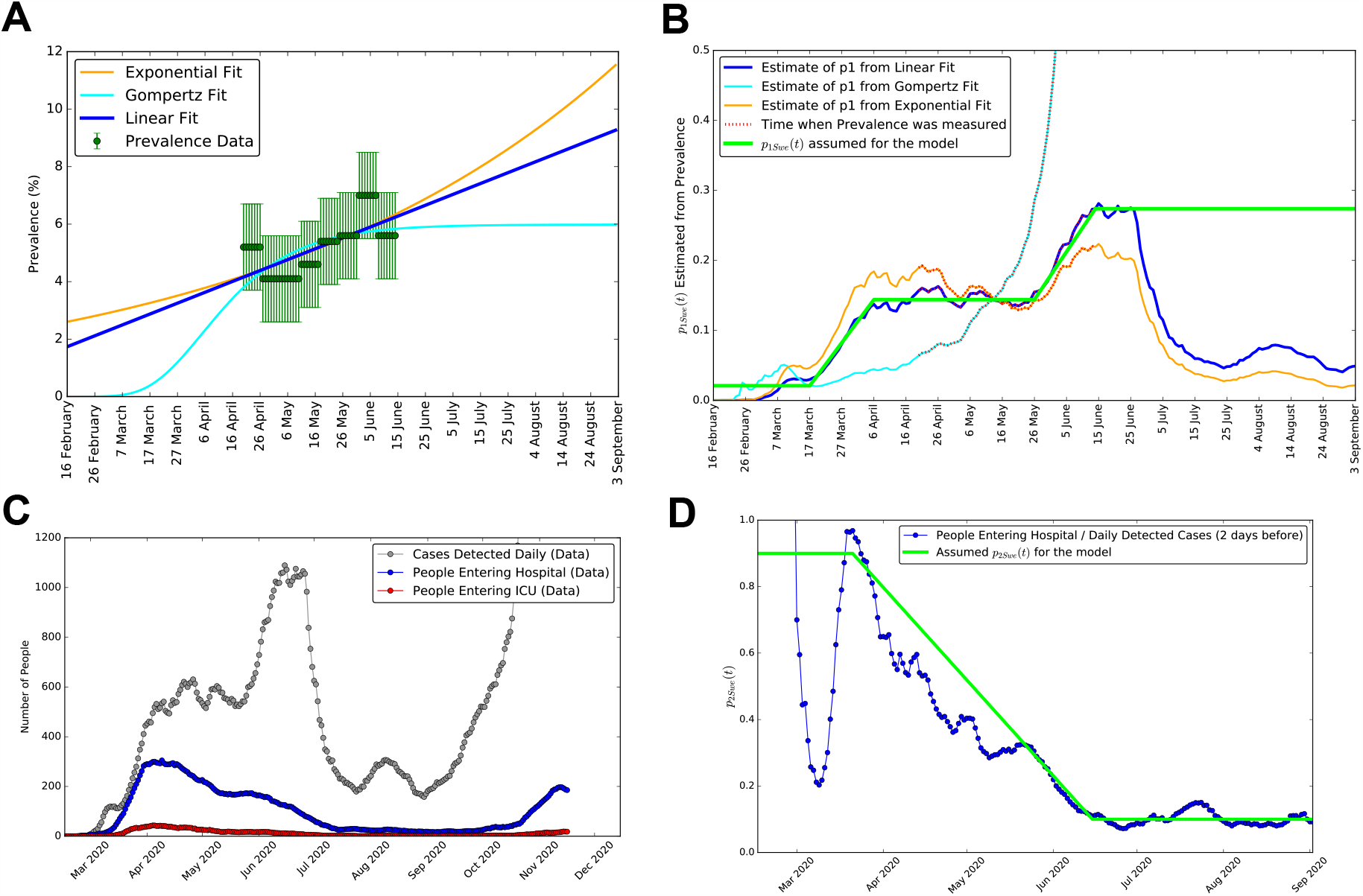
Derivation of the time-dependent probabilities of detection *p*_1*Swe*_ (*t*) and hospitalization *p*_2*Swe*_ (*t*), for Sweden. **A**: prevalence data for Sweden from [25, 26], with three alternative fits: a linear, a Gompertz and an exponential. **B**: number of detected cases divided by number of total cases inferred based on prevalence, as a function of time, for the three fits to prevalence of panel **A**. In green we report our final functional form for *p*_1*Swe*_ (*t*), i.e. the probability of being detected if infected. **C**: data for daily new detected cases, people entering hospital and people entering ICU (not to be confused with the numbers of people occupying hospitals and ICUs displayed in Fig. 2). **D**: in blue the ratio of people entering hospitals divided by new daily detected cases two days earlier, which we employ to build the piece-wise linear function in green which we assume for *p*_2*Swe*_ (*t*), i.e. the probability of being hospitalized when detected positive.

The prevalence represents the total percentage of the country’s population that is estimated to have contracted the virus, including detected and the undetected individuals. We thus can obtain an estimate of *p*_1*Swe*_ (*t*) as the ratio between the measured detected cases and the total cases estimated from the prevalence. We do so in Fig. S1, panel **B**, which reports the number of detected cases divided by the number of total cases inferred based on prevalence, as a function of time, for the three fits to prevalence of panel **A**. In green we report our final assumption for the functional form for *p*_1*Swe*_ (*t*), i.e. the probability of being detected if infected, which for Sweden we needed to be time-dependent. We assumed *p*_1*Swe*_ (*t*) to be piece-wise linear, approximately following the behaviour of the linear fit to prevalence, and saturating after the last available data point (as we do not want to introduce further assumptions). We stress here that, while the particular functional form chosen for *p*_1*Swe*_ (*t*) is somewhat arbitrary, and alternative functional shapes could have been considered, for Sweden a time-dependent *p*_1*Swe*_ (*t*) increasing with time seems indeed necessary in order for the model to fit the available data, which was not the case with Luxembourg and Austria, where *p*_1_ and *p*_2_ values constant within each wave were sufficient.

The second observation we need to incorporate in the model is that in the early phases of the pandemic in Sweden the majority of the detected cases were people needing hospitalization. This can be clearly seen from Fig. S1, panel **C**, which shows data for daily new detected cases, people entering hospital and people entering ICU (not to be confused with the number of people currently occupying hospitals and ICUs). The data of people entering hospital and ICU are publicly available on [27] and ^11^. From them, we can clearly see that in March and April the daily number of people entering hospital was more than half of the daily number of detected cases. We plot this ratio on panel **D**, reporting in blue the ratio of people entering hospitals divided by new daily detected cases two days earlier. We utilize this to build the piece-wise linear function in green which we assume for *P*_2*Swe*_ (*t*), i.e. the probability of being hospitalized when detected positive. This seemed to be between approximately 50% and 100% during March and April, as can be seen by both panels **C** and **D**, meaning the the majority of the cases that were detected positive in the early weeks/months of the epidemic in Sweden were those that required hospital treatment. This clearly changed in May and June, likely due to the change in testing strategy ordered by the government on May the 3rd [28].

### S6 Manual calibration of model parameters

We determine the values of the model parameters such that the model simulations provide a good fit to the available data, while having reasonable values w.r.t. the literature. We first in this section explain the manual calibration which we employ to get reasonable values, later on cross-validated with Markov Chain Monte Carlo methods.

The following methodology was applied similarly for Luxembourg and Austria, and to Sweden with some differences described in Sec. S5.4 and Sec. S5.3. We recall that most of the parameters have values ∈ [0; 1] due to their interpretation as rates or probabilities. The only exception is *β*, for which we considered values between 0 and 2, thus assuming a minimum average time of half a day. We initially set up *α* and *β* following literature values [49, 50]. When possible, we chose for the other parameters a tentative initial value based on domain knowledge. For example, average length of stay in hospitals or ICUs observed [48] are typically of a considerable number of days, leading to correspondingly small initial values of the rates for exiting these compartments. For the remaining parameters, e.g. those concerning non-hospitalized individuals, an initial educated guess was made based on the information available and what seemed reasonable at the time.

Next, we manually tuned the parameter values in order for the simulated model output to fit the available data. It must be stressed that, due to the model structure, individuals can only flow in one direction in the model, i.e. from being initially in the susceptible compartment, to eventually end up in either one of the recovered or dead compartments. Due to this structure, several of the parameters only influence the variables appearing downstream in the flow, not upstream. For instance, while *α, β* and *ρ* influence all compartments, the probabilities of being hospitalised or the probability of entering ICU do not influence the total detected cases. Thus, we start the manual fit by fixing those parameters that impact on the total detected cases, so that the simulated curve fits the moving average of the available data. Unlike all the others, the parameter *ρ* changes value every time that a new measure is implemented. Each value is thus fixed in order to improve the model fit to the data of total detected cases (averaged over a week) until the next change, before moving to the next value. Eventually, parameters downstream in the flow of individuals are also changed in order for the model to fit the data of the corresponding boxes. In particular, first parameters impacting total cases, then parameters impacting hospitalisations, than parameters impacting ICU and eventually those impacting death. The procedure was repeated until a satisfactory fit was achieved. The final values of parameters from our manual fit, for each country and wave of infection, are reported in Tab. S6.

**Table S6:** Parameter values for manual fit of each wave and country. All rates (*α, β* and each *τ*) are expressed in *days*^−1^, the other parameters are dimensionless. The symbol “-” means the parameter is not changed w.r.t. the value to its left in the table.

| Parameter | Manual fit Luxembourg |  |  | Manual fit Austria |  | Manual fit Sweden |  |
| --- | --- | --- | --- | --- | --- | --- | --- |
|  | 1st wave | 2nd | 3rd | 1st wave | 2nd | 1st wave | 2nd |
| $\beta$ | 1.287 | - | - | 1.287 | - | 1.287 | - |
| $\alpha$ | 0.4433 | - | - | 0.4433 | - | 0.4433 | - |
| $\tau_1$ | 0.6808 | - | - | 0.6808 | - | 0.6808 | - |
| $\tau_2$ | 0.5979 | - | - | 0.5979 | - | 0.5979 | - |
| $\tau_3$ | 0.5246 | - | - | 0.5246 | 0.5246 | 0.5246 | 0.5246 |
| $\tau_4$ | 0.0513 | 0.1050 | 0.106 | 0.0991 | 0.0948 | 0.1875 | 0.0933 |
| $\tau_5$ | 0.0617 | 0.1514 | 0.1605 | 0.1047 | 0.1240 | 0.2383 | 0.1247 |
| $\tau_6$ | 0.0853 | 0.1514 | 0.1590 | 0.1073 | 0.0856 | 0.1093 | 0.0838 |
| $\tau_7$ | 0.1084 | 0.1084 | - | 0.1084 | 0.1084 | 0.1084 | 0.1084 |
| $p_1$ | 0.31 | 0.41 | - | 0.41245 | - | $p_{1,Sw}(t)$ | 0.27389 |
| $p_2$ | 0.1534 | 0.08 | 0.042 | 0.162 | 0.065 | $p_{2,Sw}(t)$ | 0.1 |
| $p_3$ | 0.7906 | 0.86 | 0.78 | 0.72 | 0.82 | 0.72 | 0.88 |
| $p_4$ | 0.7906 | 0.9048 | 0.8868 | 0.6972 | 0.8345 | 0.6667 | 0.8965 |
| $p_5$ | 0.8717 | 0.9814 | 0.8692 | 0.8499 | 0.9275 | 0.9021 | 0.9820 |
| $p_6$ | 0.9072 | 0.9816 | 0.8868 | 0.8292 | 0.8946 | 0.7866 | 0.9731 |
| $p_7$ | 0.9876 | 0.9979 | 0.997 | 0.99 | 0.992 | 0.99 | 0.992 |

We performed this procedure for Luxembourg and for the first wave, which we conventionally assume to end with the minimum of the moving average of new daily cases occurring on June 4. It is here crucial to stress that, with the parameter set employed to fit Luxembourg simulations to data for the first wave, the model simulations once fit to the daily detected cases of the second wave lead to a significant over-prediction of the number of people in hospital and ICU for the second wave, with respect to what is observed in the data. Thus, some of the parameter values need to be changed in order for the model to fit the data of the second wave. Similarly for the third wave. For both, we repeat the procedure of manual parameter calibration described above starting from the values of the parameters for the previous wave, and incorporating any further domain knowledge about average length of stay in hospitals and so on that might have become available in the meantime.

It has to be stressed that most parameters were initially tuned for Luxembourg, and then employed for Austria and Sweden with changes in all those parameters that needed to be changed to achieve a reasonably good fit to the data, or where literature was available reporting a country-specific estimate, as it is the case for the probability of being detected *p*_1_, inferred from country-specific prevalence studies as detailed below. We further repeat the same procedure for the other countries we investigate. We consider as starting parameter values those that were employed for Luxembourg, and we further build upon that. This procedure is repeated for Austria with only changes in parameter values. However, this is not sufficient to fit Sweden. For Sweden, we further need to derive time-dependent probabilities of detection *p*_1,*Swe*_ (*t*) and of hospitalizations *p*_2,*Swe*_ (*t*), as described in detail in Sec. S5.4. these two functions are depicted in Fig. S1.

Similarly to Luxembourg, we assume the 1st wave to end with the minimum of the moving average of new daily cases on June 12 for Austria and on August 31 for Sweden. We further assume the 3rd wave in Luxembourg to start from the re-opening of schools on September the 15th. We do not split Austria or Sweden data in a third wave as only the data for Luxembourg show clearly three waves of infections, see Fig. 2. The final values of parameters from our manual fit, for each country and wave of infection, are reported in Tab. S6.

A key parameter for each country is the probability of being detected if infected, which has been derived from prevalence studies. For Luxembourg we consider this probability to be *p*_1_ = 0.31 in the first wave thanks to the Con-Vince study [46], and we assume it to be 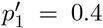 for the second and third wave based on internal communication. These numbers correspond to values for total cases over detected cases of respectively 1/0.31=3.2 and 1/0.4=2.5, which are close to the value 2.3 reported as an approximated estimate for several countries by [51]. For Austria, *p*_1_ ≈ 0.41 has been as well derived from prevalence data^12^. For Sweden we describe how we obtain *p*_1,*Swe*_ (*t*) and *p*_2,*Swe*_ (*t*) in Sec. S5.4. This estimate is also based on the prevalence data from [25, 26] displayed in Fig. S1, panel **A**. As we depict in Fig. S1, panel **B**, we assume a time-dependent *p*_1*Swe*_ (*t*) which increases from an estimated value of 0.02 in March to an estimated value of 0.27 from June onward.

### S7 Bayesian inference for cross-validation of parameters and evaluation of uncertainties

The parameter sets obtained by manual calibration are used in the manuscript to illustrate a number of qualitative and quantitative results. Nevertheless, the next question naturally arising is “how unique is each of these parameter sets, given the available data?”. We thus want to investigate to what extent the available data constrain the parameters of the model, and to which extent these parameter values can change, i.e. to quantify the uncertainties in the estimation of these parameters, given the data. A Bayesian framework provides a natural perspective from which to explore this uncertainty. For this purpose, we apply Bayesian Inference, and in particular Markov Chain Monte Carlo (MCMC) methods to I) quantify the uncertainty (via credible intervals) of the estimates of each parameter given the data, and to II) assess uncertainties on combinations of parameters (credible regions), often better identifiable than the corresponding individual parameters. In order to apply Bayesian inference and MCMC methods, we employ the dedicated python library pymcmcstat [52], version 1.9.0 [53].

#### S7.1 Sum of square residuals for the likelihood function

To perform Bayesian inference by MCMC, we need to construct a likelihood function of parameter values given the available data and to provide prior probability distributions for each parameter, which allows us to incorporate our prior knowledge. The vector of parameters, which will be specific for each country and wave, is here indicated as 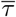.

The pymcmcstat package employs for the Likelihood 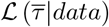 function the sum of square residuals (SSR), which is a measure of the distance between model simulation and data, such that the minimum of the SSR corresponds to the maximum of the Likelihood. This choice of the likelihood function corresponds to assume that deviations of data from the model are due to Gaussian errors, which is the simplest assumption to make when we have no additional knowledge of the potential sources of errors.

The SSR for Luxembourg reads:

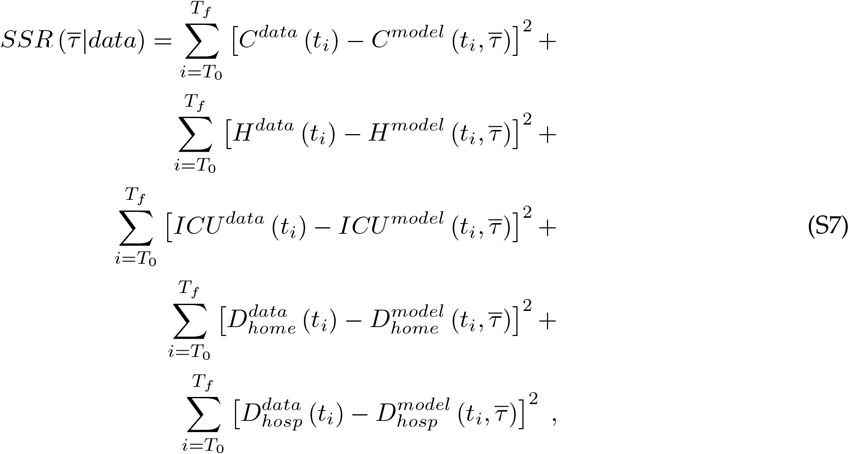

where 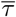 is the array of parameters, *t*_*i*_ = *T*_0_, *T*_0_ + 1, …, *T*_*f*_ 1, *T*_*f*_ indicates the number of days from the beginning of the epidemic (assumed for that country), *T*_0_ and *T*_*f*_ are respectively the first and last day of the wave under investigation (1st wave, 2nd wave or, for Luxembourg, 3rd wave), *C* is the cumulative total number of detected cases, *H* is the number of people in hospital, *ICU* is the number of people in ICU, *D*_*hosp*_ and *D*_*home*_ are respectively the number of people dead in hospital (including ICU) or outside hospital (*D*_*home*_ includes also nursing houses). The same SSR was employed for Austria and Sweden (with the corresponding data and model variables), except that the two compartments for dead were merged into one compartment due to lack of more fine-grained data.

The quantities in Eq. S7 derived from the model, omitting the dependencies on time and parameters for ease of notation, in terms of the model variables (Tab. S1) are given by:

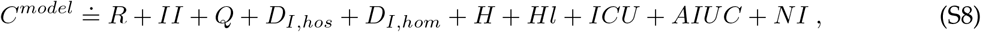

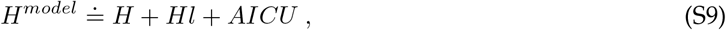

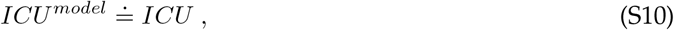

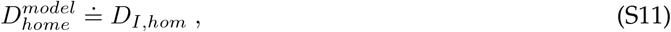

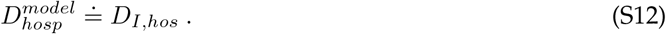

#### S7.2 Prior probability distributions

The goal of our Bayesian inference approach is to assess, given the model structure and the public data which we consider in this work, which are the constraints that these data impose on the parameter values, i.e. how much freedom we would have in selecting alternative parameter sets. This is a way to assess the uncertainties on the estimates of these parameters and the uncertainties on combination of parameters, sometimes better identified than the individual parameters. For the Bayesian inference approach, for each country and wave we assume flat prior probability distributions on each parameter over a parameter space as large as possible, within reason. The reasons in doing so are mainly two. First, flat priors are the simplest choice which does not require any additional knowledge except the intervals over which these priors should be extended. Second, flat priors are the less informative choice we can make, thus we are not introducing any bias toward a particular parameter set. We choose not to introduce any bias by means of the priors because we want to use this approach to cross-validate the manually calibrated parameter set, thus we only want the result to depend on the public time-series data, and on a reasonable limitation of the available parameter space by means of the priors.

We select a flat prior probability distribution which for each step of the social interaction parameter *ρ*_*n*_ ranges from 0 to 1. For each rate, it ranges from 0 days^−1^ to 1 days^−1^, except for beta where it ranges from 0 days^−1^ to 2 days^−1^, and for each probability from 0 to 0.5 or 0.5 to 1 depending what the probability represents. We opt for these somewhat reduced parameter space w.r.t. the interval from 0 to 1 because, without loss of generality (we do not expect probabilities of dying being above 0.5, i.e. 50%, and so on), it is less demanding in terms of time and computational resources needed for the MCMC chains to converge. Similarly, we further restricted the priors between 0 and 0.5 for those rates associated to an average length of stay which is known to be considerably larger than a day, e.g. for the rate of exiting ICU. These flat prior probability distributions are reported for each country and wave as the light gray area in Fig. S2, where we can appreciate the extent to which the posterior probability distributions are shrunk w.r.t. the priors. The posteriors are depicted via their 50% and 90% credible regions, respectively in blue and cyan.

#### S7.3 Markov Chain Monte Carlo (MCMC) method

After defining the likelihood function and the priors, we calculated the posterior probability distribution over the parameter space (for each country and wave separately) by means of Markov Chain Monte Carlo (MCMC) methods available via the dedicated python library pymcmcstat.

Among the different Metropolis based sampling techniques provided by pymcmcstat, we employ the Delayed Rejection Adaptive Metropolis (DRAM) algorithm. It is a combination of the Delayed-Rejection (DR) algorithm, which delays rejection by sampling from a narrower distribution, and the Adaptive-Metropolis (AM) algorithm, which adapts the covariance matrix of the proposal Gaussian distribution at specified intervals.

In order to increase the speed of our sampling, we run in parallel 8 chains (except for Sweden, where 6 chains for the first wave and 4 chains for the second were run, due to limited available computational resources). Each chain is initialized at random initial conditions extracted from the flat prior probability distribution for each parameter, which ensures that no bias is introduced by the choice of the chains’ initial conditions. As we will see in the next section, having multiple chains will also be useful to assess convergence of the chains. We run each chain for 500000 iterations, as such high number of iterations appeared to ensure chain convergence, as measured by the method described in the next sections. The first half of each chain is automatically discarded as burn-in (default settings of the package, which were kept), while the second half is employed to determine the posterior probability distributions.

#### S7.4 Thinning of the chains only for visualization

Thinning of chains (only considering one sample every several) is not usually appropriate when the goal is precision of estimates from an MCMC sample [54]. Thus, for the estimate of parameters or credible intervals we did not employ thinning. Nevertheless, the same study points out that thinning can be useful for other reasons, such as memory or time constraints in post-chain processing. We employed thinning for the purpose of generating the figures containing estimates of the posteriors, e.g. Fig. S3. For generating this type of figures, we thinned the chains keeping only every 100th sample.

The same study [54] suggests that a better course of action than thinning, however, is to generate multiple independent chains. It suggests monitoring different independent chains and assaying whether the estimates produced by the different chains are suitably similar to each other. This is performed, for example, when implementing the Gelman-Rubin diagnostic [55, 56] to consider the variation among these independent chains. We thus apply this procedure.

#### S7.5 Ensuring convergence of MCMC chains through Gelman-Rubin diagnostic

In our case, across countries and waves, visual inspection of the posterior probability distributions estimates derived by each chain independently (a simple approach also performed in [13]) reveals that for most parameters it appears as though all the chains have converged to about the same distribution (with the exception of very few parameters where one or few chains lead to somewhat different posteriors). We nevertheless further employed a more solid approach.

Determining whether or not MCMC chains have converged to the posterior density is a significant challenge when performing MCMC simulations. There are many diagnostics available for assessing chain convergence. As stressed by [54], a robust approach is to use the Gelman-Rubin diagnostic [55, 56], which requires several sets of chains for comparison. The Gelman-Rubin approach essentially performs an analysis of the variances within each chain set and between each chain set. The same diagnostic has been employed for the same purpose in the framework of MCMC convergence in modelling COVID-19 in e.g. [12, 14].

When we compute from the full chains the Gelman-Rubin diagnostics, we indeed obtain values of R, the so-called “Potential Scale Reduction Factor (PSRF)”, that are extremely close to 1 for most parameters. In general, R close to 1 indicates the chains have converged [55, 56].

### S8 Results from the MCMC

We cross-validated the parameter sets we employed by means of Bayesian inference and Markov Chain Monte Carlo methods, as described in Sec. S7. In order to confirm the viability of the choice of parameter values from the manual calibration, we show in Sec. 3.1 that the simulations generated by our model fit the available data for these countries. Additionally, we show in Sec. S8.1 that the manually calibrated sets of parameters which we employ for the simulations are consistent with the Markov Chain Monte Carlo estimates, which in turn shows that the available data constrain parameters only to some extent, leaving considerable uncertainties. We based our MCMC solely on publicly available time-series data, which results in wide uncertainties. With the manually calibrated parameter set, we had to some extent overcome these uncertainties by employing parameter values obtained from literature estimates based on data other than the publicly available time-series, which we refer to as domain knowledge. Examples are the estimates of length of stay of patients in ICUs and hospitals. We finally observe in Sec. S8.2 that there is degeneracy between parameters, with combinations (e.g. ratios) of parameters being better constrained by the data than individual parameters.

#### S8.1 Manually calibrated parameter sets compatible with Bayesian inference estimate, which underlines wide uncertainties in parameter values

The parameter sets from the manual calibration of the model discussed in Sec. S6 are summarized in Tab. S6, Tab. S3, Tab. S4 and Tab. S5 and are employed through the manuscript. Fig. S2 shows the Bayesian estimates of these parameters given the time series data, obtained by MCMC (credible intervals) and employed to cross-validate them.

Posteriors are indicated by their 50% (blue) and 90% (cyan) credible intervals, and we can appreciate that for the majority of parameters, these posteriors are considerably shrunk w.r.t. the flat prior probability distributions we assumed for every parameter (in Sec. S7.2). Nevertheless, for some parameters this is not the case and we obtained posteriors which are almost flat and wide as the priors, which means those parameters are not well identified based on the available data and model structure. We also report in Fig. S2 point-estimates from the MCMC, i.e. the maximum a posteriori estimate, and the mean a posteriori estimate of each parameter. These simply are, respectively, the maximum and the mean of the posterior for each parameter. For rather symmetric distributions the two values tend to correspond, while for very skewed distributions they happen to be very different. Moreover, for almost flat posterior distributions, like e.g. those for most of the parameters *τ*_*i*_ with *i* = 4, 5, 6, the maximum is not that representative of the distribution.

In Fig. S2 we see that, for all countries and waves, the values of the manual fit (green) mostly are very close to either the maximum or the mean a posteriori estimate. It is most of the times within the 50% credible interval of the posterior, or at least inside the 90% credible interval, with only a couple of exceptions. Thus, we see here that our manually calibrated sets of parameters are fully consistent with our Bayesian estimate based on MCMC.

However, we can also appreciate that for each of those parameters, credible intervals are relatively wide, which indicates that the uncertainties on the estimate of these parameters based on the data alone are rather large. In particular we see, at a qualitative level, that the values of the social interaction parameter *ρ*_*n*_ seem to be in general better constrained than the other parameters, with the probabilities *p*_*i*_ being slightly better constrained than the rates *τ*_*i*_, in general very poorly identified except for *α* and *β* which are affected by smaller uncertainties w.r.t. other rates. We also observe the probability *p*_7_ to be extremely well constrained (to values close to 1) in most of the waves and countries.

#### S8.2 Quantifying uncertainties on estimate of parameters by MCMC: wide uncertainties, but degeneracy in parameter estimations

The domain of the posterior probability distribution obtained via the MCMC, Sec. S7, has the same number of dimensions as the number of parameters considered, which for instance is 19 for the 1st wave of Luxembourg. When we are interested in one parameter at a time, we project the chains on one dimension at a time (which, if we had an analytic form for the posterior, would be done by integrating over all parameters except the one of interest, thus obtaining the marginal posterior distribution). This is how we obtained the 1D posteriors displayed in Fig. S2. The next question that arises naturally is if the wide uncertainties affecting some parameters are an effect of having projected to a lower dimensional space, or if certain parameters are really so poorly identified as it seems. We will see that both types of situations arise, depending on the parameter.

To investigate this, we thus project the posterior distribution over two parameters at a time, and we repeat this for each couple of parameters in Fig. S3 for the MCMC run of the 1st Wave of Luxembourg, and in Fig. S4, S5, S6, S7, S8 and S9 for the other waves and countries. We can clearly see from the above mentioned figures that, while certain parameter pairs are very well constrained, others are not and we have parameter pairs that appear to be for instance correlated or anti-correlated. In general we observe that there is a lot of degeneracy between parameters. This means that there is not a unique combination of parameters that allow a good fit of the model to the data, but many of them.

We moreover observe combinations (e.g. ratios) of parameters being better constrained than the individual parameters. Consider e.g. the parameters *α*, the inverse of the mean incubation period, and *β*, the mean reproduction rate, which are in common with a standard SEIR model. Their credible intervals are displayed in Fig. S2. When we consider the joint posterior over the two parameters, which is visible close to the center of Fig. S3 for Luxembourg, Fig. S6 for Austria and Fig. S8 for Sweden, we remark that this posterior is narrow and rotated toward the diagonal direction, showing anti-correlation between the estimates of the two parameters, meaning that low values of *β* can only be considered together with high values of *α*, and the other way around. These patterns occur for other parameters as well, and they are conserved to some extent across countries, as they occur with the same properties for the 1st wave of Austria in Fig. S8, and to some extent for Sweden, so they are likely specific rather to the model’s structure and parameters than to the country. We also observe other interesting patterns, e.g. anti-correlation between each *ρ*_*n*_ and the subsequent in time, clearly visible in the 2nd and 3rd waves of Luxembourg, the 2nd wave of Sweden and to some extent in the 1st and 2nd waves of Austria. This can easily be understood in terms of the model: lowering the social interaction one time-period and increasing it the next, or doing the converse, can equally lead to the same good agreement with data.

Very interestingly, while certain couples of parameters seem to be well constrained, at least when considered two at a time, for other parameters these projected posteriors are still close to flat, resulting in squares that are predominantly red in Fig. S3. This effect is weaker for certain waves and countries, with the 3rd wave of Luxembourg and the 2nd wave of Sweden providing the more narrow constraints on parameters. This nevertheless is stronger for others, being the strongest in the 1st wave of Sweden, where parameters seem to be very poorly identified. This could also be related to the dimension of the domain of the posterior (the higher the number of parameters, the less constrained), as the 3rd wave of Luxembourg and the 2nd wave of Sweden both have indeed “only” 14 parameters each, while the 1st wave of Sweden has 26 of them.

**Fig. S2.**
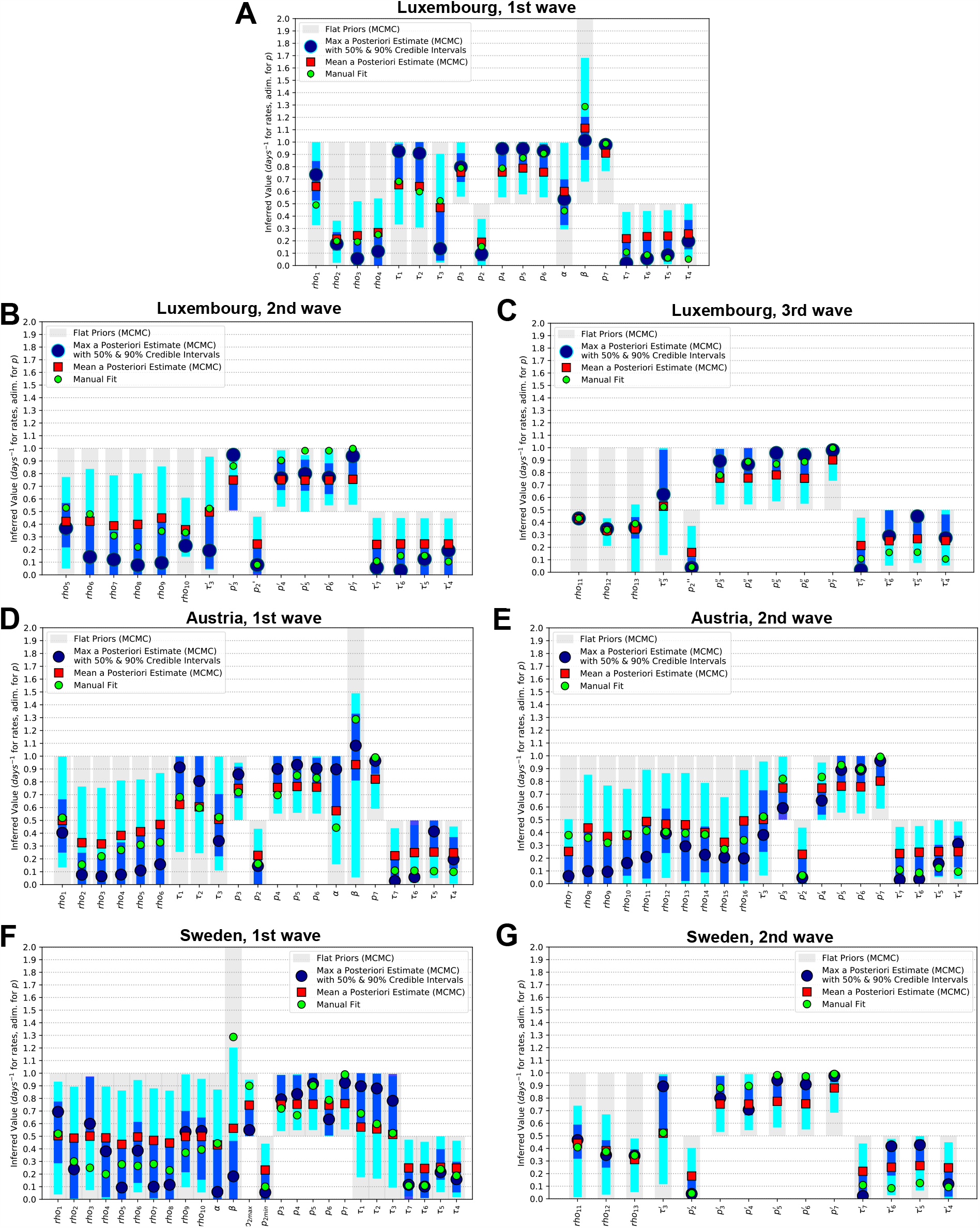
Posterior probability distributions from MCMC for each parameter, compared with priors and with manual fits. Each country and wave. Luxembourg (**A**: 1st wave, **B**: 2nd wave, **C**: 3rd wave), Austria (**D**: 1st wave, **E**: 2nd wave) and Sweden (**F**: 1st wave, **G**: 2nd wave). For each parameter, prior probability distributions are reported as gray areas, while the posteriors are reported by means of their 50% (blue) and 90% (cyan) credible intervals, with their maxima reported as large blue dots and their means as red squares. The parameter values for the manual fit (Tab. S6 for probabilities and rates, Tab. S3, Tab. S4 and Tab. S5 for *ρ*_*n*_) are reported as green small circles.

**Fig. S3.**
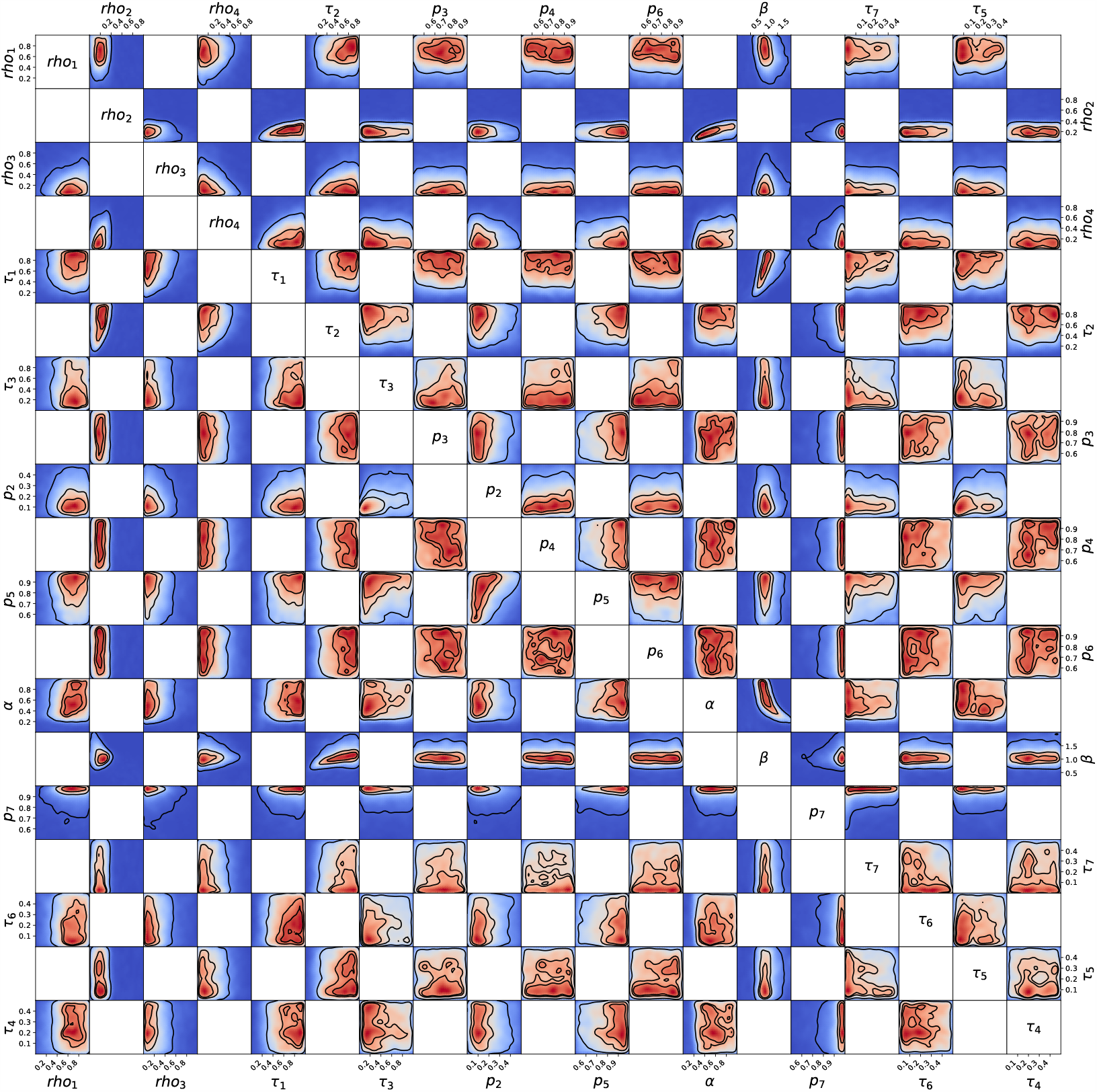
Posterior probability distribution from MCMC projected over each pair of parameters, for the 1st wave of Luxembourg. 2D projections of the posterior probability distribution estimated as described in Sec. S7, corresponding to the 1D projections of Fig. S2. Each square represents the 2D projection of the posterior over two parameters. The parameter name and scale are reported only at the margins of the figure and on the diagonal, for visual clarity, but they apply for all subplots. As the figure would be symmetric along the diagonal, each projection is only reported once, leading to the white squares. The heatmap colors represent posterior probability, with red being high and blue being low, thus the true values of the parameters have a higher probability to be in regions colored in red than in regions colored in blue, with dark blue representing a probability close to 0. The three black contours reported in each panel represent respectively the 25%, 50% and 90% Bayesian credible regions. Correlations, anti-correlations and more complex degeneracy between parameter pairs are visible. The same figures for other waves and countries are available in the Supplementary Material as Fig. S4 for the 2nd wave of Luxembourg, Fig. S5 for the 3rd wave of Luxembourg, Fig. S6 for the 1st wave of Austria, Fig. S7 for the 2nd wave of Austria, Fig. S8 for the 1st wave of Sweden, Fig. S9 for the 2nd wave of Sweden.

**Fig. S4.**
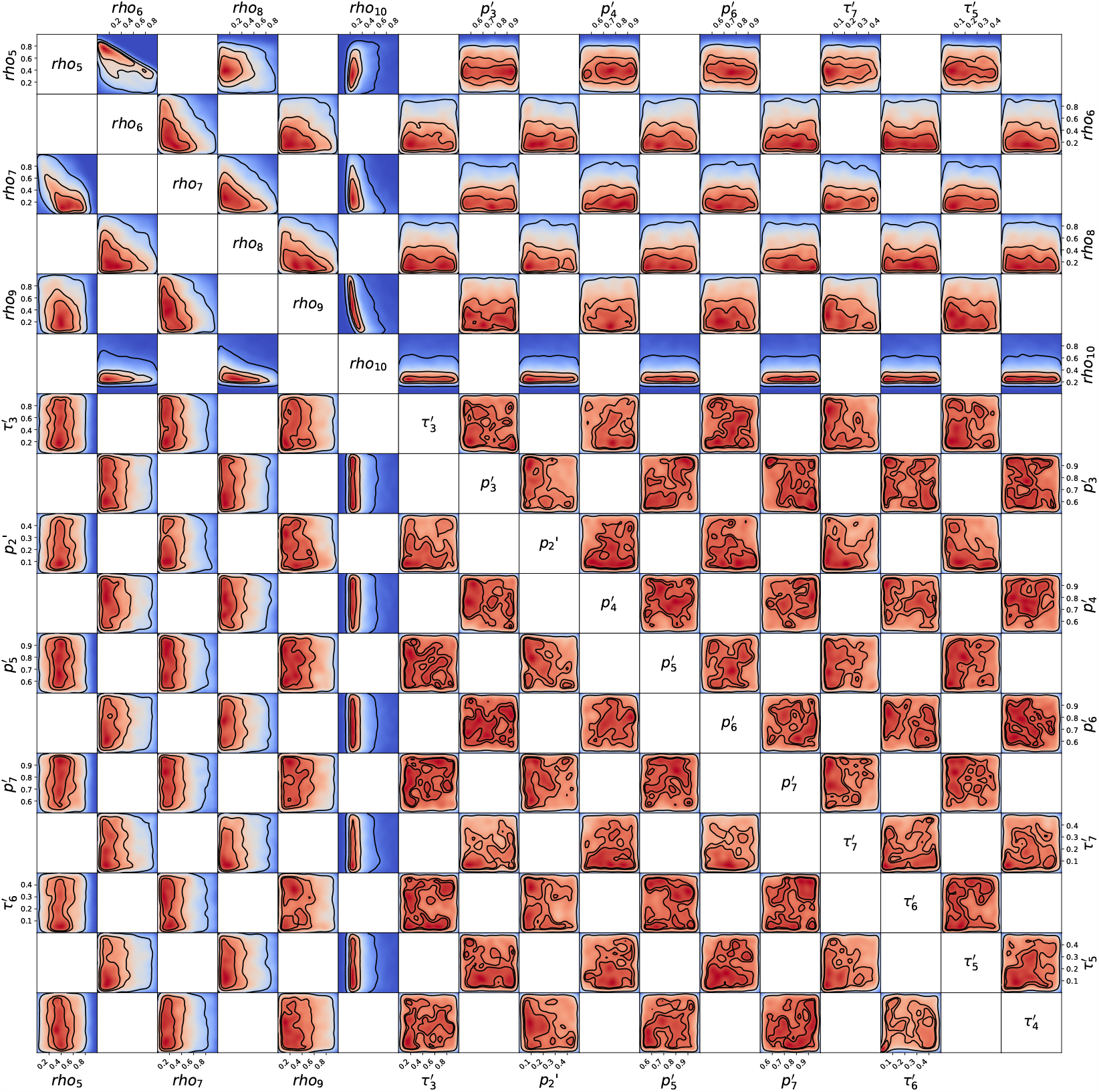
Posterior probability distribution from MCMC projected over each couple of parameters, for the 2nd wave of Luxembourg. 2D projections of the posterior probability distribution estimated as described in Sec. S7, corresponding to the 1D projections of Fig. S2. Each square represents the 2D projection of the posterior over two parameters. The parameter name and scale are reported only at the margins of the figure and on the diagonal, for visual clarity, but they apply for all subplots. As the figure would be symmetric along the diagonal, each projection is only reported once, leading to the white squares. The heatmap colors represent posterior probability, with red being high and blue being low, thus the true values of the parameters have a higher probability to be in regions colored in red than in regions colored in blue, with dark blue representing a probability close to 0. The three black contours reported in each subpanel represent respectively the 25%, 50% and 90% Bayesian credible regions. Correlations, anti-correlations and more complex degeneracy between couples of parameters are visible.

**Fig. S5.**
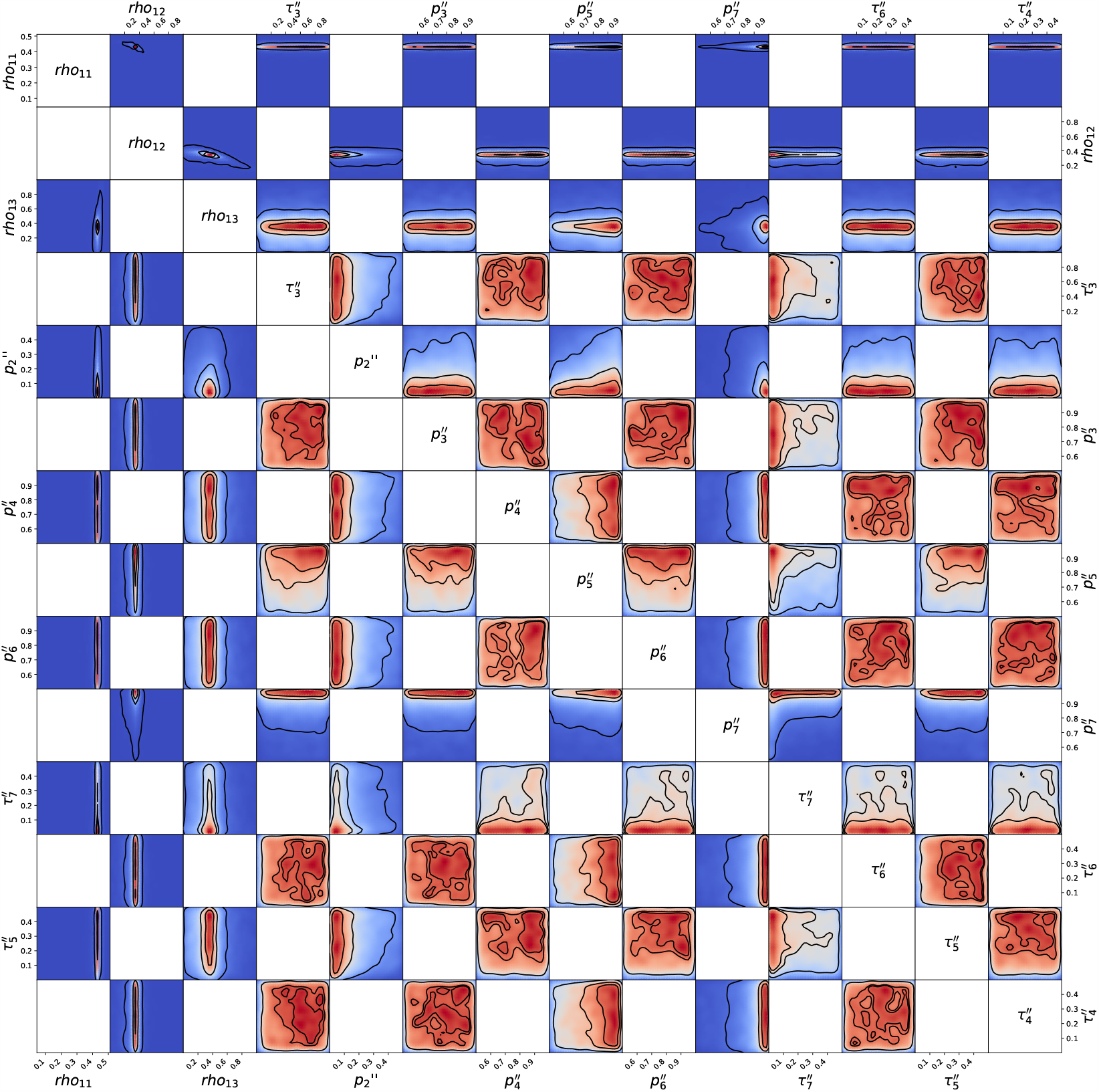
Posterior probability distribution from MCMC projected over each couple of parameters, for the 3rd wave of Luxembourg. 2D projections of the posterior probability distribution estimated as described in Sec. S7, corresponding to the 1D projections of Fig. S2. Each square represents the 2D projection of the posterior over two parameters. The parameter name and scale are reported only at the margins of the figure and on the diagonal, for visual clarity, but they apply for all subplots. As the figure would be symmetric along the diagonal, each projection is only reported once, leading to the white squares. The heatmap colors represent posterior probability, with red being high and blue being low, thus the true values of the parameters have a higher probability to be in regions colored in red than in regions colored in blue, with dark blue representing a probability close to 0. The three black contours reported in each subpanel represent respectively the 25%, 50% and 90% Bayesian credible regions. Correlations, anti-correlations and more complex degeneracy between couples of parameters are visible.

**Fig. S6.**
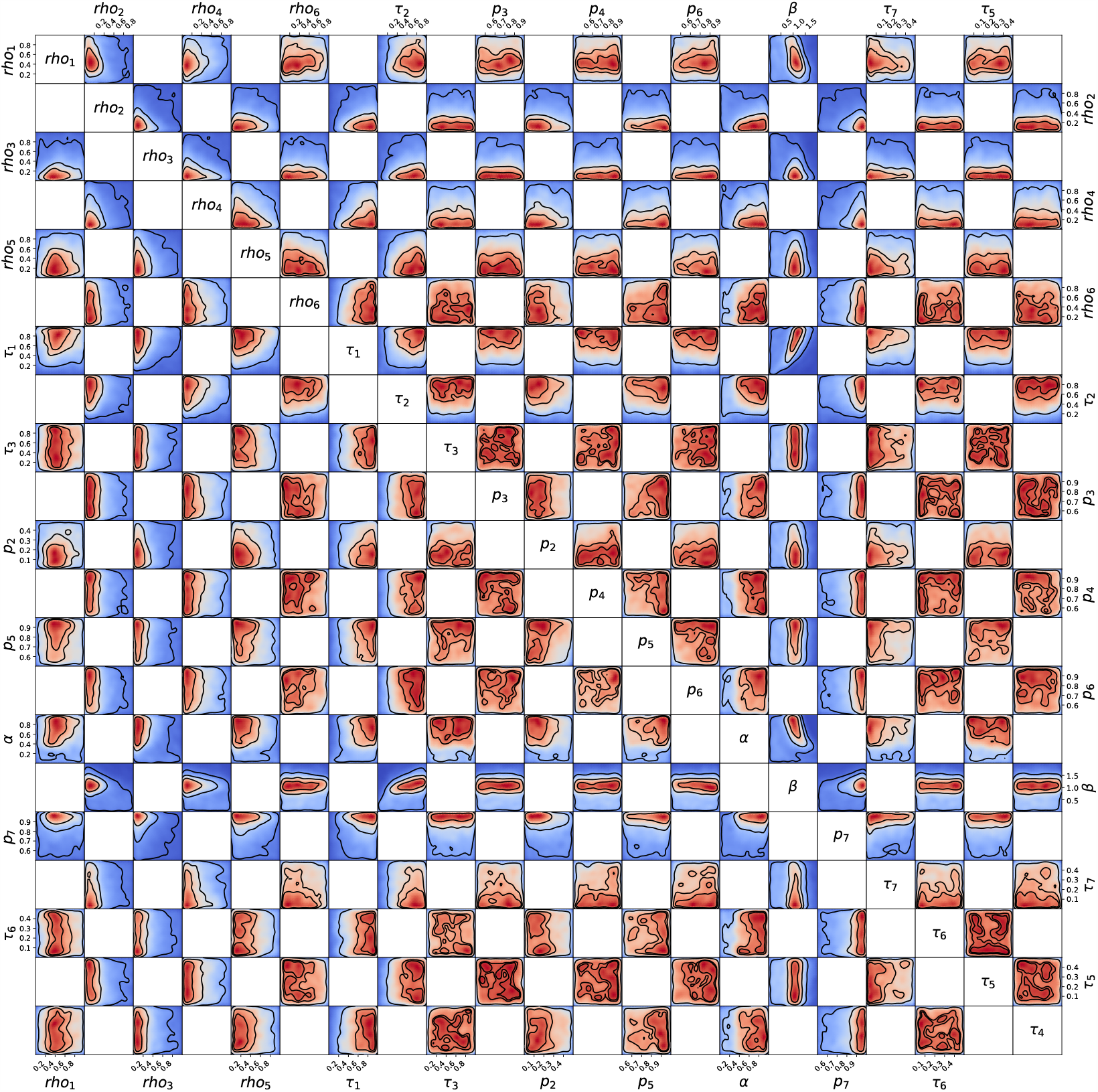
Posterior probability distribution from MCMC projected over each couple of parameters, for the 1st wave of Austria. 2D projections of the posterior probability distribution estimated as described in Sec. S7, corresponding to the 1D projections of Fig. S2. Each square represents the 2D projection of the posterior over two parameters. The parameter name and scale are reported only at the margins of the figure and on the diagonal, for visual clarity, but they apply for all subplots. As the figure would be symmetric along the diagonal, each projection is only reported once, leading to the white squares. The heatmap colors represent posterior probability, with red being high and blue being low, thus the true values of the parameters have a higher probability to be in regions colored in red than in regions colored in blue, with dark blue representing a probability close to 0. The three black contours reported in each subpanel represent respectively the 25%, 50% and 90% Bayesian credible regions. Correlations, anti-correlations and more complex degeneracy between couples of parameters are visible.

**Fig. S7.**
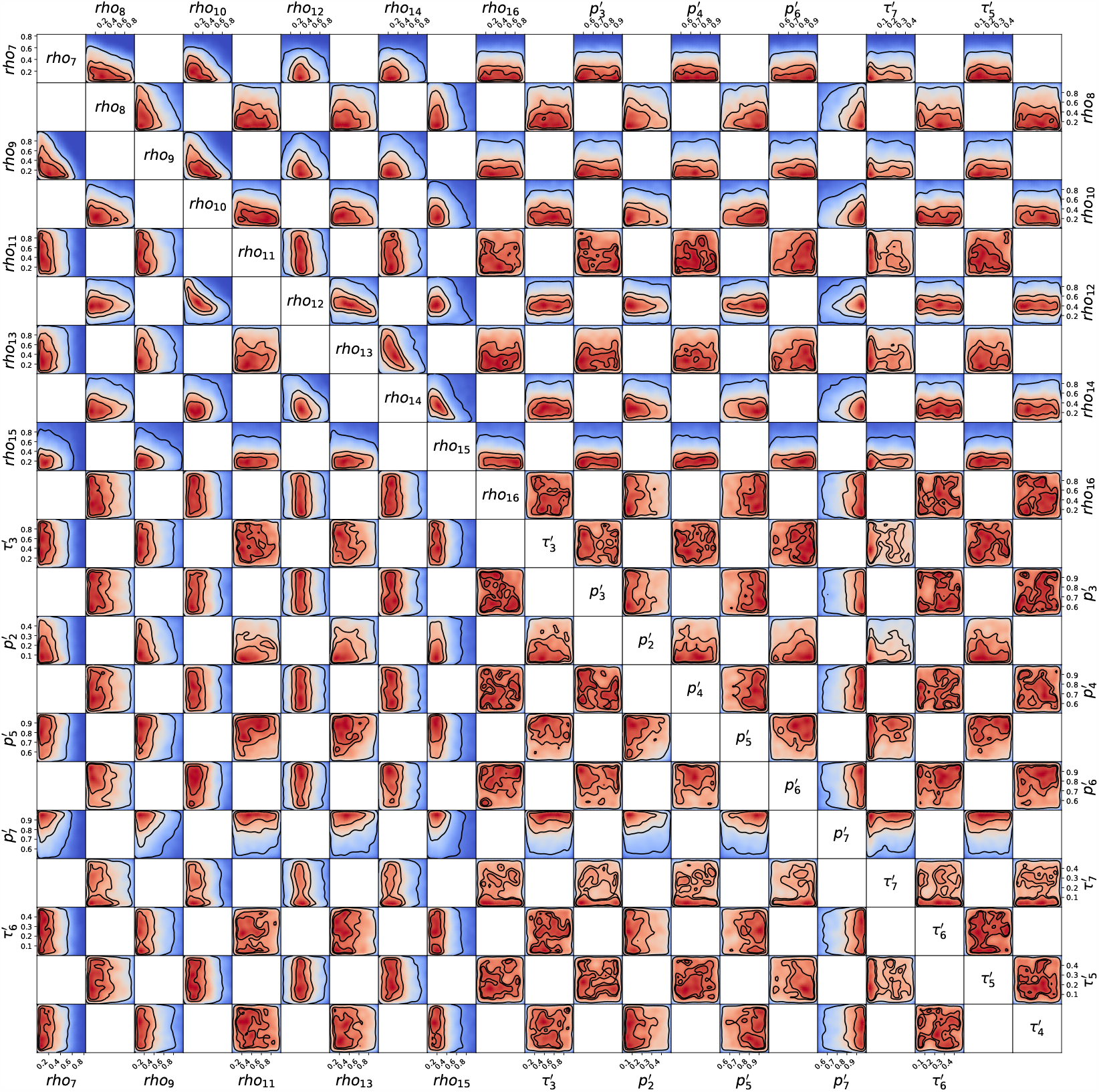
Posterior probability distribution from MCMC projected over each couple of parameters, for the 2nd wave of Austria. 2D projections of the posterior probability distribution estimated as described in Sec. S7, corresponding to the 1D projections of Fig. S2. Each square represents the 2D projection of the posterior over two parameters. The parameter name and scale are reported only at the margins of the figure and on the diagonal, for visual clarity, but they apply for all subplots. As the figure would be symmetric along the diagonal, each projection is only reported once, leading to the white squares. The heatmap colors represent posterior probability, with red being high and blue being low, thus the true values of the parameters have a higher probability to be in regions colored in red than in regions colored in blue, with dark blue representing a probability close to 0. The three black contours reported in each subpanel represent respectively the 25%, 50% and 90% Bayesian credible regions. Correlations, anti-correlations and more complex degeneracy between couples of parameters are visible.

**Fig. S8.**
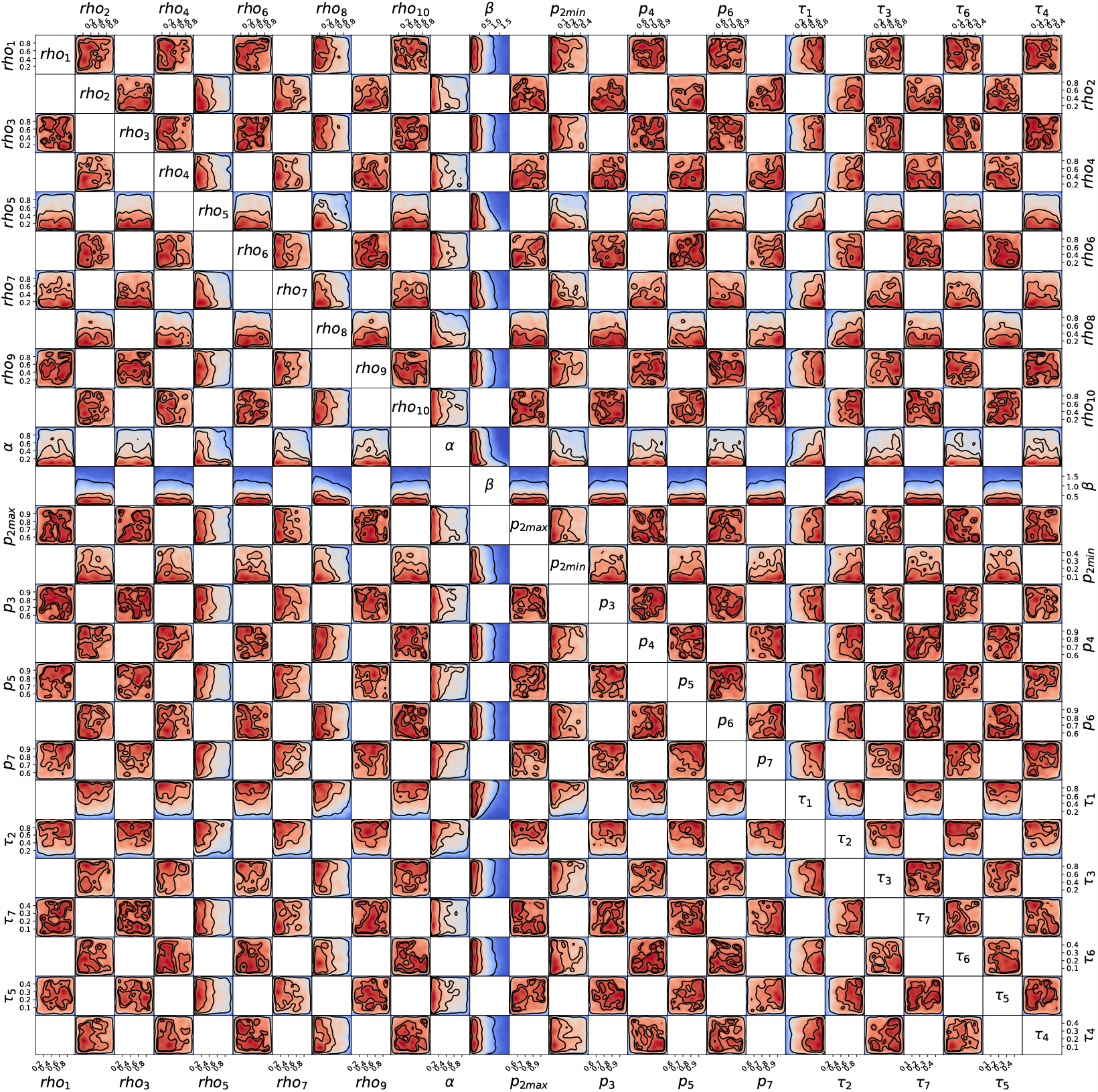
Posterior probability distribution from MCMC projected over each couple of parameters, for the 1st wave of Sweden. 2D projections of the posterior probability distribution estimated as described in Sec. S7, corresponding to the 1D projections of Fig. S2. Each square represents the 2D projection of the posterior over two parameters. The parameter name and scale are reported only at the margins of the figure and on the diagonal, for visual clarity, but they apply for all subplots. As the figure would be symmetric along the diagonal, each projection is only reported once, leading to the white squares. The heatmap colors represent posterior probability, with red being high and blue being low, thus the true values of the parameters have a higher probability to be in regions colored in red than in regions colored in blue, with dark blue representing a probability close to 0. The three black contours reported in each subpanel represent respectively the 25%, 50% and 90% Bayesian credible regions. Correlations, anti-correlations and more complex degeneracy between couples of parameters are visible.

**Fig. S9.**
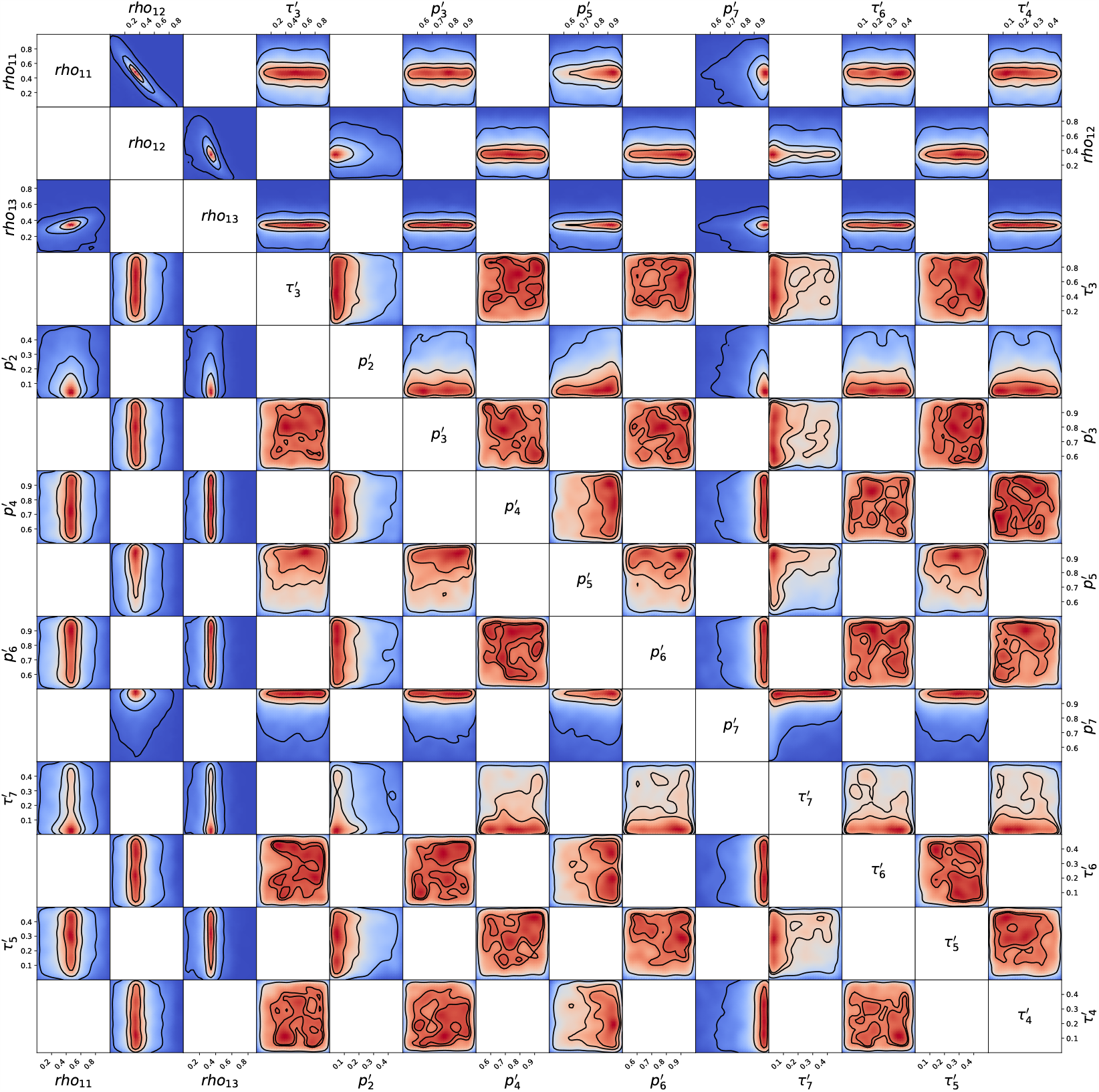
Posterior probability distribution from MCMC projected over each couple of parameters, for the 2nd wave of Sweden. 2D projections of the posterior probability distribution estimated as described in Sec. S7, corresponding to the 1D projections of Fig. S2. Each square represents the 2D projection of the posterior over two parameters. The parameter name and scale are reported only at the margins of the figure and on the diagonal, for visual clarity, but they apply for all subplots. As the figure would be symmetric along the diagonal, each projection is only reported once, leading to the white squares. The heatmap colors represent posterior probability, with red being high and blue being low, thus the true values of the parameters have a higher probability to be in regions colored in red than in regions colored in blue, with dark blue representing a probability close to 0. The three black contours reported in each subpanel represent respectively the 25%, 50% and 90% Bayesian credible regions. Correlations, anti-correlations and more complex degeneracy between couples of parameters are visible.

## Footnotes

1 https://www.icuregswe.org/en/data--results/COVID-19-in-swedish-intensive-care/

2 https://covid19.public.lu/fr/graph.html

3 https://www.acaps.org/covid19-government-measures-dataset, public independent database recognised by the WHO, last accessed on 03/12/2020.

4 https://covid19.public.lu/fr/mesures-sanitaires-en-vigueur.html, last accessed 15/12/2020.

5 https://covid19.public.lu/fr/graph.html, last accessed on 15/12/2020.

6 https://de.wikipedia.org/wiki/COVID-19-Pandemie_in_\unhbox\voidb@x\bgroup\accent127O\protect\penalty\@M\hskip\z@skip\egroupsterreich (collection of various sources), accessed on 15/12/2020.

7 https://covid19-dashboard.ages.at/dashboard_Hosp.html, accessed on 15/12/2020.

8 https://www.sora.at/nc/news-presse/news/news-einzelansicht/news/COVID-19-praevalenz-1006. html, accessed on 15/12/2020.

9 https://www.krisinformation.se/en/news, accessed on 15/12/2020.

10 https://www.folkhalsomyndigheten.se/smittskydd-beredskap/utbrott/aktuella-utbrott/covid-19/statistik-och-analyser/bekraftade-fall-i-sverige/, https://www.icuregswe.org/en/data--results/covid-19-in-swedish-intensive-care/, https://c19.se, accessed on 15/12/2020.

11 https://www.icuregswe.org/en/data--results/COVID-19-in-swedish-intensive-care/

12 https://www.sora.at/nc/news-presse/news/news-einzelansicht/news/covid-19-praevalenz-1006. html, accessed on 15/12/2020.

